# Effective tree-based classification for automated flow cytometry data analysis on samples with suspected haematological malignancy

**DOI:** 10.1101/2022.12.07.22283209

**Authors:** Alex Rothwell, Anthony Carter, Peter Green, Andrew R Jones

## Abstract

Flow cytometry is a commonly used diagnostic technique for haematological malignancies. The gold standard method for analysis of flow cytometry data is manual gating, which is time consuming and requires a highly skilled operator, generating a bottleneck in the workflow and potentially increasing time to diagnose malignancy. For nearly 20 years attempts have been made at replacing manual analysis with automated algorithms, however these are not deemed accurate enough for clinical practice. Clustering methods have been the focus of previous automated attempts, though supervised methods have been shown to be more accurate and require less manual intervention. Tree-based classification algorithms make decisions using an analogous process to manual gating. One hundred and fifty-two flow cytometry files were generated from peripheral blood samples of patients with suspected haematological malignancies. A trained operator labelled events in these files as one of nine cell types. CART, Random Forest and XGBoost were trained on the labelled dataset and the performance was evaluated against previously published clustering methods. Classification algorithms showed higher mean F1 scores than clustering methods. There was no significant difference between CART, Random Forest and XGBoost mean F1 scores, and all three algorithms showed mean prediction times per sample of less than 25 seconds. Tree-based methods struggled to differentiate B cell subtypes, which show similar phenotypic signatures and present an area for future improvement. This work demonstrates the effectiveness of tree-based classification algorithms for flow cytometry analysis. Overall, CART may offer a solution to automated flow cytometry analysis for the purpose of haematological malignancies due to showing high agreement with manual analysis, and short prediction and training times.

## Introduction

Around 327,800 people in the UK, and 3.1 million people worldwide, have been diagnosed with cancer-related haematological malignancies [1,2]. Flow cytometry (FC) is a commonly used diagnostic technique for such disorders [3]. Cells from a liquid sample (such as blood) are stained with fluorochrome-conjugated antibodies which bind to surface and intracellular markers. The markers and/ or cells of interest will determine the antibody panel used for staining. Cells are sequentially passed through a laser and fluorescent light signals of the excited fluorochromes are measured, as well as the side and front scatter which indicate cell size and internal complexity, respectively. Marker expression is proportional to the measured emitted photons once the variable emission profiles of the applied fluorescent dyes are considered [4]. This allows cells to be identified based on marker expression, for example, it is known the majority of normal B cells express CD45, CD19 and CD20, though in Chronic lymphocytic leukaemia (CLL), malignant B cells additionally express CD5 and CD23 [5,6]. This allows for the identification of malignant cell populations, enabling diagnosis and monitoring of disease [4,7,8].

The gold standard FC data analysis technique for the diagnosis of haematological malignancies is a process called “manual gating”. Manual gating involves visual inspection of “events”, individual records of each cell, viewed in one- or two-dimensional plots and the partitioning of known events into sub-populations in a hierarchical manner by drawing “gates” using a software interface [8]. Summary statistics are then reported, such as the percentage that the identified sub-populations make up of a wider population, the absolute counts of events in a sub-population, and the median fluorescence intensity (MFI) of a fluorochrome of a sub-population [9]. This allows for the formation of a composite phenotype of a sample, based on the pattern of antigen expression, which can be compared to World Health Organization classification guidance to indicate disease type [10]. This report, as well as the reports generated from other diagnostic tests, will be examined by a clinician who will form a diagnosis [7]. Manual gating is an inefficient, time-consuming process, with the operators’ bias potentially influencing results [11,12]. The process creates a bottleneck in the FC workflow and potentially increases the time it takes to diagnose malignancy [13]. Yet, early detection of cancer is likely the most effective strategy for reducing mortality rates [14]. Therefore, automated analysis of FC data for the diagnosis of haematological malignancies, which has the potential to be faster and less biased than current strategies, could provide both medical and economic benefit.

For nearly 20 years, a plethora of attempts have been made at replacing manual gating systems with automated algorithms [15,16]. However, these have not become widely adopted in clinical practice, with a recent survey indicating that 80% of FC clinical laboratories worldwide “never” or “rarely” use automated FC analysis software and only 2% “usually” use it [17]. The key reason being that even the most advanced automated systems currently available fail to meet several needs of clinical laboratories, not least, the systems are not deemed accurate enough [18].

Some successful attempts have been made at classifying FC samples by diagnosis, where the model input is the FC data generated from a single patient’s sample and the output is a diagnosis for the patient [19–22]. However, methods which classify each event within an FC sample, where the model input is the FC data from a single event and the output is the label of that event e.g. “B cell”, likely fit into the current diagnostic pipeline more effectively, reducing barriers to adoption and increasing the likelihood of these methods being integrated in clinical practice. This is because, although performing diagnosis may be straightforward based on the results of FC analysis with some analysis software even suggesting a diagnosis, performing diagnosis is the duty of clinicians, not the laboratory staff who perform FC testing [23,24].

Past attempts at automated FC analysis have focused on the use of clustering algorithms to group similar events into sub-populations [15]. Some of these attempts have gained substantial interest and have been subject to several benchmarking and comparison studies [8,25]. There are several advantages to clustering approaches. Firstly, labelling a dataset to be used for training classification algorithms requires knowledge of the identity of each event within a sample, whereas clustering can be used to identify novel sub-populations which were previously unknown [17]. Secondly, clustering algorithms can be applied to any FC sample, regardless of how data was obtained. In contrast, classification methods typically require the unseen data to possess the same input features the model was trained using, meaning that a trained classifier could only be used on unseen data generated using the same antibody panel, reducing its utility [26]. Furthermore, the labelling of datasets required as training data for classification algorithms is time consuming and expensive [27]

Despite their somewhat limited use in the field thus far, classification algorithms which label each event in a sample possess properties which may make them an attractive proposition for use within clinical practice. Firstly, trained classification algorithms have been demonstrated to be more accurate on FC data than clustering algorithms which are entirely unsupervised [8]. Secondly, the primary aim of FC analysis for the diagnosis of haematological malignancy is to identify known populations. Identifying unknown, novel populations, which clustering algorithms are well suited for, is not the goal [28]. Furthermore, clustering approaches still require some manual intervention, as sub-populations require manual labelling, somewhat negating potential benefits of automated analysis.

CART (classification and regression trees), Random Forest (RF) and XGBoost (XGB) are tree-based algorithms which can be used for classification. CART are binary trees which mathematically determine the best split on single features. Training these trees involves repeatedly performing these splits, partitioning the prediction space based on simple rules. Predicting the class of an unseen data instance requires following the series of rules generated by the tree to determine a classification. Each rule follows the form of testing whether a feature is higher or lower than a value e.g. CD45 expression > 0.5 [29]. RFs create an ensemble of trees, each tree is varied due to having been limited to searching over a random subset of features on a random sample of training data when generating decision rules, with the output being the class voted by most trees [30]. XGB is a gradient boosting algorithm. It combines a series of weak tree classifiers which sequentially aim to further minimise the training error of the previous tree, ultimately resulting in a strong classifier [31]. RF and XGB are less likely to overfit training data than a single decision tree [32]. Tree-based classifiers could be particularly well suited to FC analysis, as the manual gating process is analogous to the decision-making process tree-based algorithms use, the label given to an event being dependent on whether fluorescence intensities are above or below certain thresholds. The aim of this study was to develop an automated event labelling approach for FC analysis which utilises tree-based classification algorithms, and test against previously published clustering methods.

## Results

### Clustering Method and Classification Algorithm Performance

The 100,000 events in 152 fcs files, generated from patient samples with suspected haematological malignancy, were first manually labelled based on cell type. This dataset was used to train and test the effectiveness of tree-based classification algorithms and previously published clustering methods at automated event labelling. Classification algorithms were trained twice, once with the models weighting training instances inversely proportional to class frequency in the dataset, and once without.

Cell clusters identified by the clustering methods were matched to the labelled populations using the Hungarian assignment algorithm which maximises the sum of F1 scores across populations, allowing no population to be matched more than once. Clustering methods showed poorer performance than classification algorithms (mean F1 score: *clustering methods;* 0.28 ± 0.2 vs *classification algorithms;* 0.94 ± 0.04) (Fig 1). Cytometree achieved the highest mean F1 score of any clustering method (0.60 ± 0.21). All other clustering methods showed F1 scores of less than 0.5 (Table 1). The tree-based classification models showed consistently high mean F1 score (CART: 0.94 ± 0.02, RF: 0.95 ± 0.05, XGB: 0.93 ± 0.02). A two-way ANOVA showed that there was not a statistically significant interaction between the classifier algorithm and weighting training samples (F (2, 54) = 1.09, p = 0.34). Simple main effects analysis showed that F1 score was not significantly different between classifier algorithms (p = 0.05), with the simpler CART models performing as well as the more complex RF and XGB models. Additionally, balancing training weights did not significantly affect F1 score (p = 0.25), indicating that balancing of minority classes was not an effective mechanism for improving overall model performance. However, balancing training weights led to the XGB models exhibiting the lowest mean precision score of any classification algorithm (0.90), and the highest mean recall (0.96), (Supplementary Table 1 and 2).

**Fig 1.**
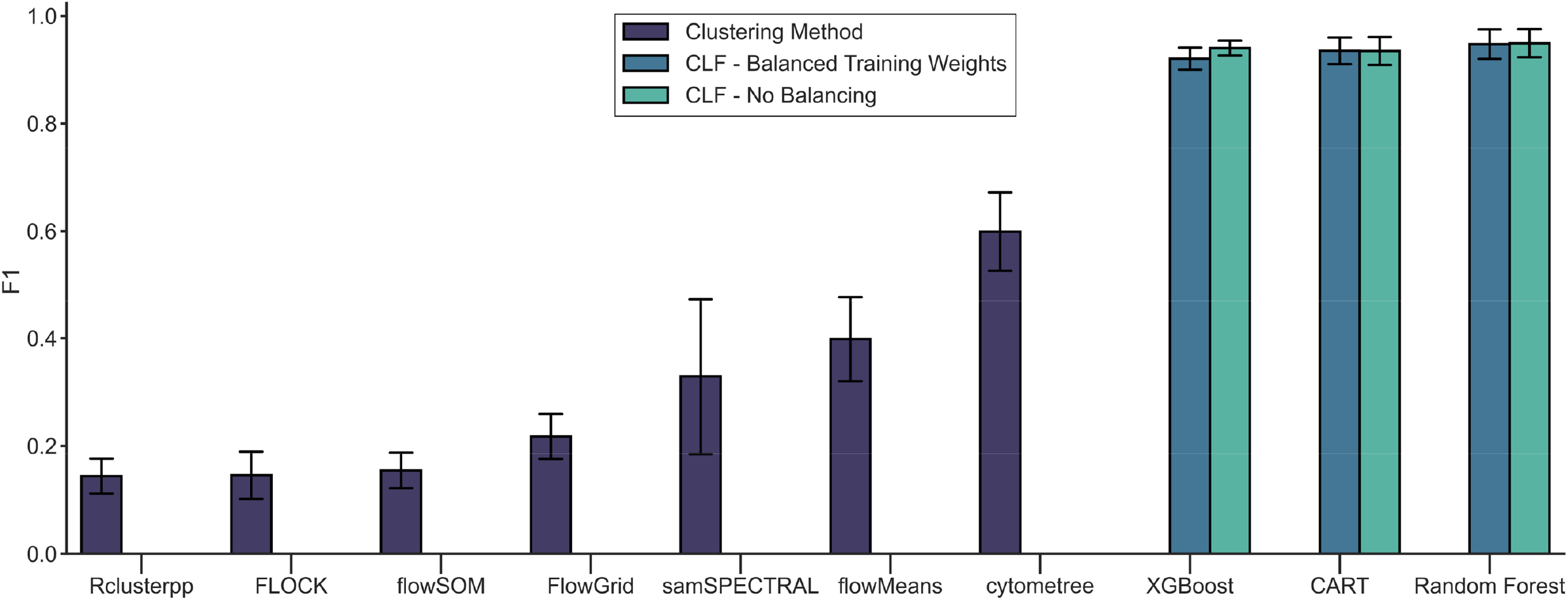
Tree-based classification algorithms exhibit higher mean F1 scores than clustering methods for automated flow cytometry analysis. Mean (± SD) macro F1 score for classification algorithms and clustering methods. Classification algorithms (CLF) were trained once with models weighting classes inversely proportional to their frequencies within the dataset during model training, and once without. Classification algorithms were cross validated over 10 folds with a labelled dataset containing events from 152 samples. Clustering methods were run on 10 randomly chosen samples from the labelled dataset, identified clusters were matched to labelled populations using the Hungarian assignment algorithm.

**Table 1.** Mean (± SD) F1 score for each class for each classification algorithm and clustering method.

| Method \ Class | CD4 + T cell | Dual + T cell | Dual – T cell | CD8 + T cell | G/D T cell | Kappa B cell | Lambda B cell | NK cell | Not Lymph | Overall |
| --- | --- | --- | --- | --- | --- | --- | --- | --- | --- | --- |
| Random Forest-No Balancing | 1.0 (± 0.0) | 0.82 (± 0.13) | 0.99 (± 0.02) | 1.0 (± 0.0) | 0.98 (± 0.01) | 0.9 (± 0.08) | 0.87 (± 0.15) | 0.99 (± 0.0) | 1.0 (± 0.0) | **0.95 (± 0.04)** |
| Random Forest-Balanced Training Weights | 1.0 (± 0.0) | 0.81 (± 0.14) | 0.99 (± 0.02) | 1.0 (± 0.0) | 0.98 (± 0.01) | 0.89 (± 0.08) | 0.87 (± 0.15) | 0.99 (± 0.01) | 1.0 (± 0.0) | **0.95 (± 0.05)** |
| XGBoost-No Balancing | 1.0 (± 0.0) | 0.8 (± 0.05) | 1.0 (± 0.0) | 1.0 (± 0.0) | 0.99 (± 0.0) | 0.87 (± 0.04) | 0.81 (± 0.06) | 0.99 (± 0.0) | 1.0 (± 0.0) | **0.94 (± 0.02)** |
| CART-Balanced Training Weights | 1.0 (± 0.0) | 0.75 (± 0.13) | 0.98 (± 0.03) | 1.0 (± 0.0) | 0.97 (± 0.02) | 0.88 (± 0.09) | 0.86 (± 0.13) | 0.99 (± 0.01) | 1.0 (± 0.0) | **0.94 (± 0.05)** |
| CART-No Balancing | 1.0 (± 0.0) | 0.76 (± 0.1) | 0.98 (± 0.04) | 1.0 (± 0.0) | 0.96 (± 0.03) | 0.88 (± 0.09) | 0.86 (± 0.12) | 0.99 (± 0.01) | 1.0 (± 0.0) | **0.94 (± 0.04)** |
| XGBoost-Balanced Training Weights | 1.0 (± 0.0) | 0.64 (± 0.12) | 1.0 (± 0.0) | 1.0 (± 0.0) | 0.99 (± 0.0) | 0.88 (± 0.04) | 0.82 (± 0.06) | 0.98 (± 0.01) | 1.0 (± 0.0) | **0.92 (± 0.03)** |
| Cytometree | 0.92 (± 0.05) | 0.12 (± 0.13) | 0.61 (± 0.37) | 0.74 (± 0.11) | 0.28 (± 0.17) | 0.7 (± 0.32) | 0.58 (± 0.34) | 0.67 (± 0.28) | 0.78 (± 0.13) | **0.6 (± 0.21)** |
| flowMeans | 0.86 (± 0.3) | 0.0 (± 0.0) | 0.1 (± 0.3) | 0.73 (± 0.38) | 0.01 (± 0.01) | 0.46 (± 0.42) | 0.2 (± 0.4) | 0.41 (± 0.37) | 0.84 (± 0.09) | **0.4 (± 0.25)** |
| samSPECTRAL | 0.35 (± 0.44) | 0.0 (± 0.01) | 0.18 (± 0.34) | 0.24 (± 0.38) | 0.28 (± 0.41) | 0.68 (± 0.35) | 0.35 (± 0.43) | 0.19 (± 0.39) | 0.67 (± 0.44) | **0.33 (± 0.35)** |
| FlowGrid | 0.48 (± 0.11) | 0.0 (± 0.0) | 0.0 (± 0.0) | 0.18 (± 0.18) | 0.01 (± 0.01) | 0.36 (± 0.42) | 0.17 (± 0.34) | 0.18 (± 0.21) | 0.58 (± 0.12) | **0.22 (± 0.15)** |
| flowSOM | 0.06 (± 0.17) | 0.0 (± 0.0) | 0.0 (± 0.0) | 0.16 (± 0.26) | 0.01 (± 0.03) | 0.36 (± 0.44) | 0.18 (± 0.37) | 0.0 (± 0.0) | 0.62 (± 0.14) | **0.15 (± 0.16)** |
| FLOCK | 0.06 (± 0.17) | 0.0 (± 0.0) | 0.0 (± 0.0) | 0.01 (± 0.03) | 0.0 (± 0.0) | 0.34 (± 0.42) | 0.17 (± 0.35) | 0.0 (± 0.0) | 0.73 (± 0.25) | **0.15 (± 0.14)** |
| Rclusterpp | 0.18 (± 0.17) | 0.0 (± 0.0) | 0.0 (± 0.0) | 0.22 (± 0.25) | 0.01 (± 0.02) | 0.29 (± 0.36) | 0.14 (± 0.29) | 0.0 (± 0.01) | 0.45 (± 0.06) | **0.14 (± 0.13)** |
| **Mean: Clustering method** | **0.41 (± 0.2)** | **0.02 (± 0.02)** | **0.13 (± 0.14)** | **0.33 (± 0.23)** | **0.08 (± 0.09)** | **0.46 (± 0.39)** | **0.26 (± 0.36)** | **0.21 (± 0.18)** | **0.66 (± 0.18)** | **0.28 (± 0.2)** |
| **Mean: Classification algorithm** | **1.0 (± 0.0)** | **0.76 (± 0.11)** | **0.99 (± 0.02)** | **1.0 (± 0.0)** | **0.98 (± 0.01)** | **0.88 (± 0.07)** | **0.85 (± 0.11)** | **0.99 (± 0.01)** | **1.0 (± 0.0)** | **0.94 (± 0.04)** |
| **Mean: All** | **0.69 (± 0.11)** | **0.37 (± 0.06)** | **0.53 (± 0.08)** | **0.64 (± 0.12)** | **0.5 (± 0.06)** | **0.66 (± 0.24)** | **0.53 (± 0.24)** | **0.57 (± 0.1)** | **0.82 (± 0.09)** | **0.59 (± 0.12)** |

### Training and Prediction Time

Training time showed greater variation between classification algorithms than F1 score (CART: 447 ± 21 seconds, RF: 7,027 ± 443 seconds, XGB: 103,285 ± 10,579 seconds). On average, XGB took 14.7 times as long to run as RF and 231 times as long to run as CART (Fig 2).

**Fig 2.**
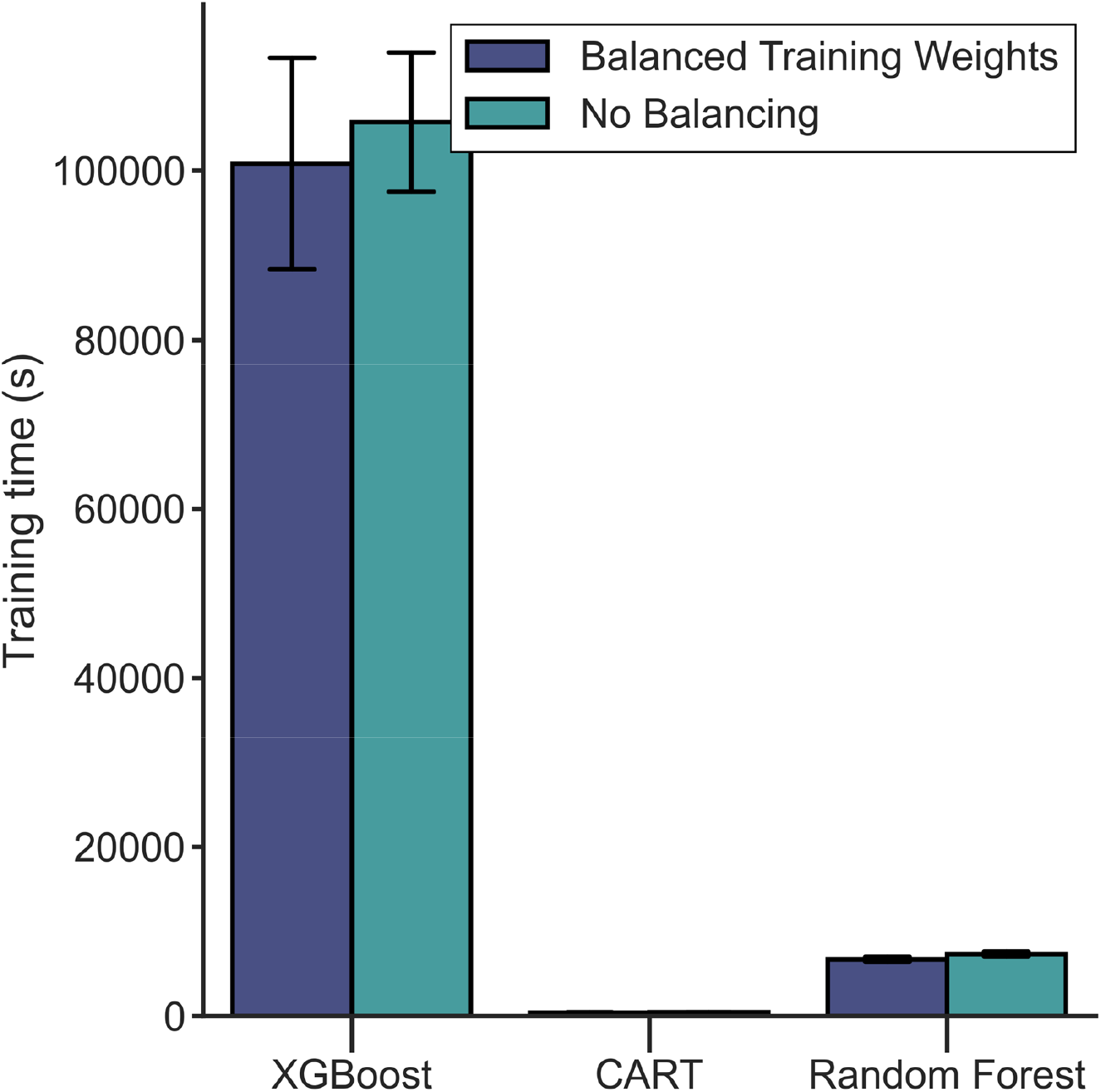
XGBoost takes considerably longer to train than CART and Random Forest. Mean (± SD) training time for classification algorithms. Classification algorithms were trained once with models weighting classes inversely proportional to their frequencies within the dataset during model training, and once without. XGBoost was trained on data from 110 flow cytometry samples containing 100,000 events, whereas CART and Random Forest were trained on data from 137 samples.

RF showed the longest mean prediction time per sample (22.69 ± 2.29 seconds) with CART and XGB taking less than 2 seconds on average (CART: 0.20 ± 0.02 seconds, XGB: 1.64 ± 0.39 seconds) (Fig 3). Cytometree took the longest mean time to make predictions of any clustering method (19.42 seconds), though this was highly variable between samples and repeats (SD: 23.82 seconds).

**Fig 3.**
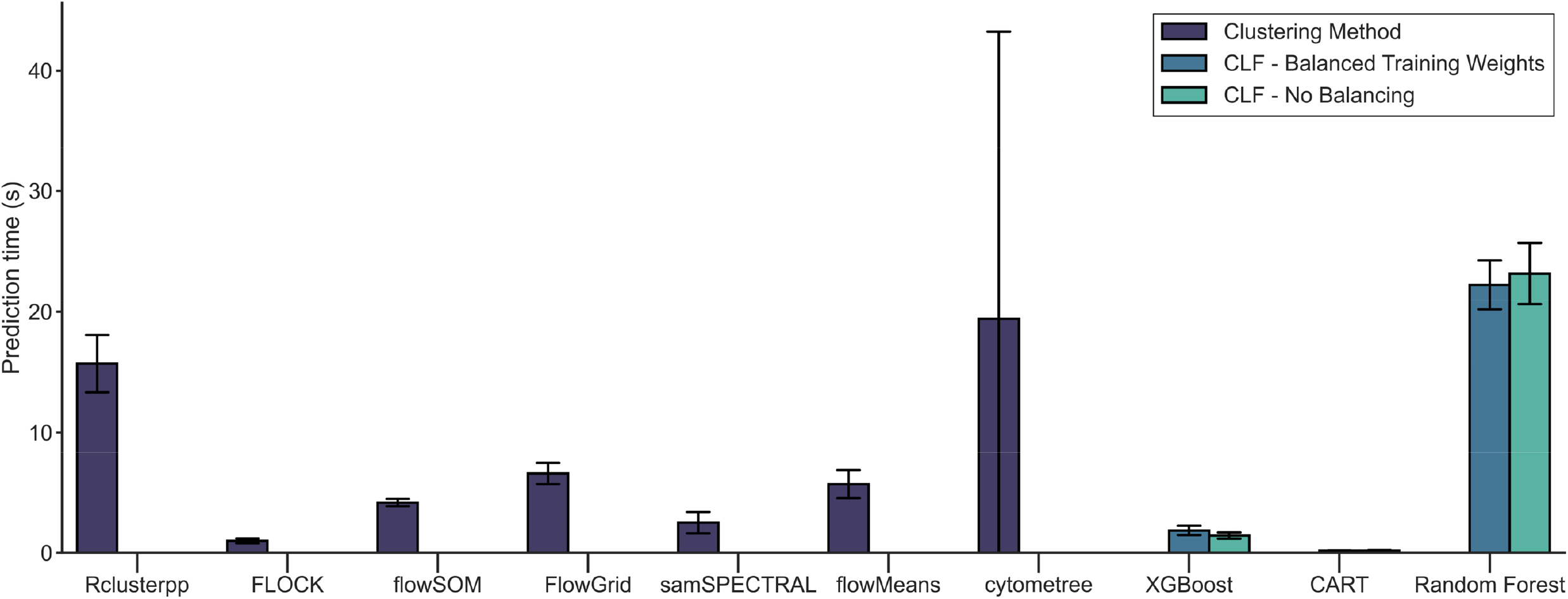
All methods showed fast mean prediction times per sample. Mean (± SD) prediction time per sample for classification algorithms and clustering methods. Classification algorithms (CLF) were trained once with models weighting classes inversely proportional to their frequencies within the dataset during model training, and once without. Classification algorithms were cross validated over 10 folds with a labelled dataset containing events from 152 samples. Clustering methods were run on 10 randomly chosen samples from the labelled dataset.

Fig 4 depicts mean prediction time and F1 score. Data points in the bottom right of the figure show the most favourable algorithms, those with fast prediction times and high F1 scores. XGB and CART models showed these characteristics, though XGB suffered from long training times. RF showed high F1 score but slower prediction time, in the top right of the plot. Data points in the bottom left of the plot showed fast prediction times and low F1 scores, FLOCK had the mean fastest prediction time of clustering methods (0.99 ± 0.20 seconds), though low mean F1 scores (0.15 ± 0.14).

**Fig 4.**
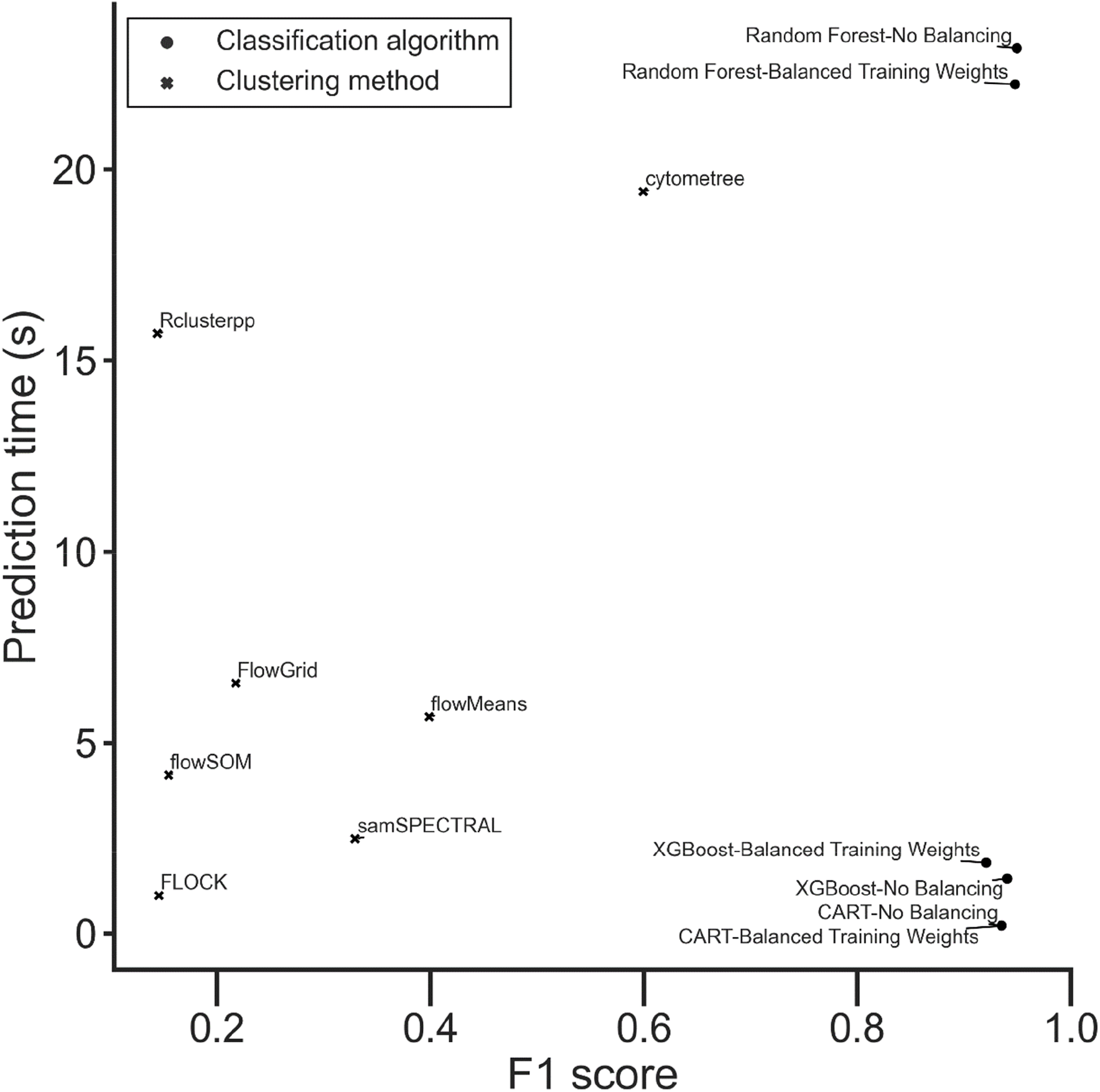
CART and XGBoost showed fast mean prediction times and high mean F1 scores. Mean macro F1 score vs mean prediction time in seconds for classification algorithms and clustering methods. Classification algorithms are shown as dots, clustering methods as crosses. Classification algorithms were trained once with models weighting classes inversely proportional to their frequencies within the dataset during model training, and once without. Classification algorithms were cross validated over 10 folds with a labelled dataset containing events from 152 samples. Clustering methods were run on 10 randomly chosen samples from the labelled dataset, identified clusters were matched to labelled populations using the Hungarian assignment algorithm.

### Individual Class Performance

Table 1 shows F1 scores within each class for each algorithm, Supplementary Information includes tables showing precision and recall for each class. Fig 5 shows aggregated confusion matrices for all classification algorithms and clustering methods. Ideal performance would be shown by a black diagonal running top-left to bottom-right, with all other cells light green colour – this would imply all cell types were 100% correctly labelled. “CD4/CD8 + T cell” (i.e. Dual + T cell) was the most challenging class to correctly predict showing the lowest mean F1 score of all classes for clustering methods (0.02 ± 0.02) and classification algorithms (0.76 ± 0.11). As shown in Fig 5, even the classification algorithms (CART, RF, XGB), which performed best, have a tendency to classify some “CD4/CD8 + T cell” samples as either “CD4 + T cell” or “CD8 + T cell” (variations in colour across the second row from the top in each panel).

**Fig 5.**
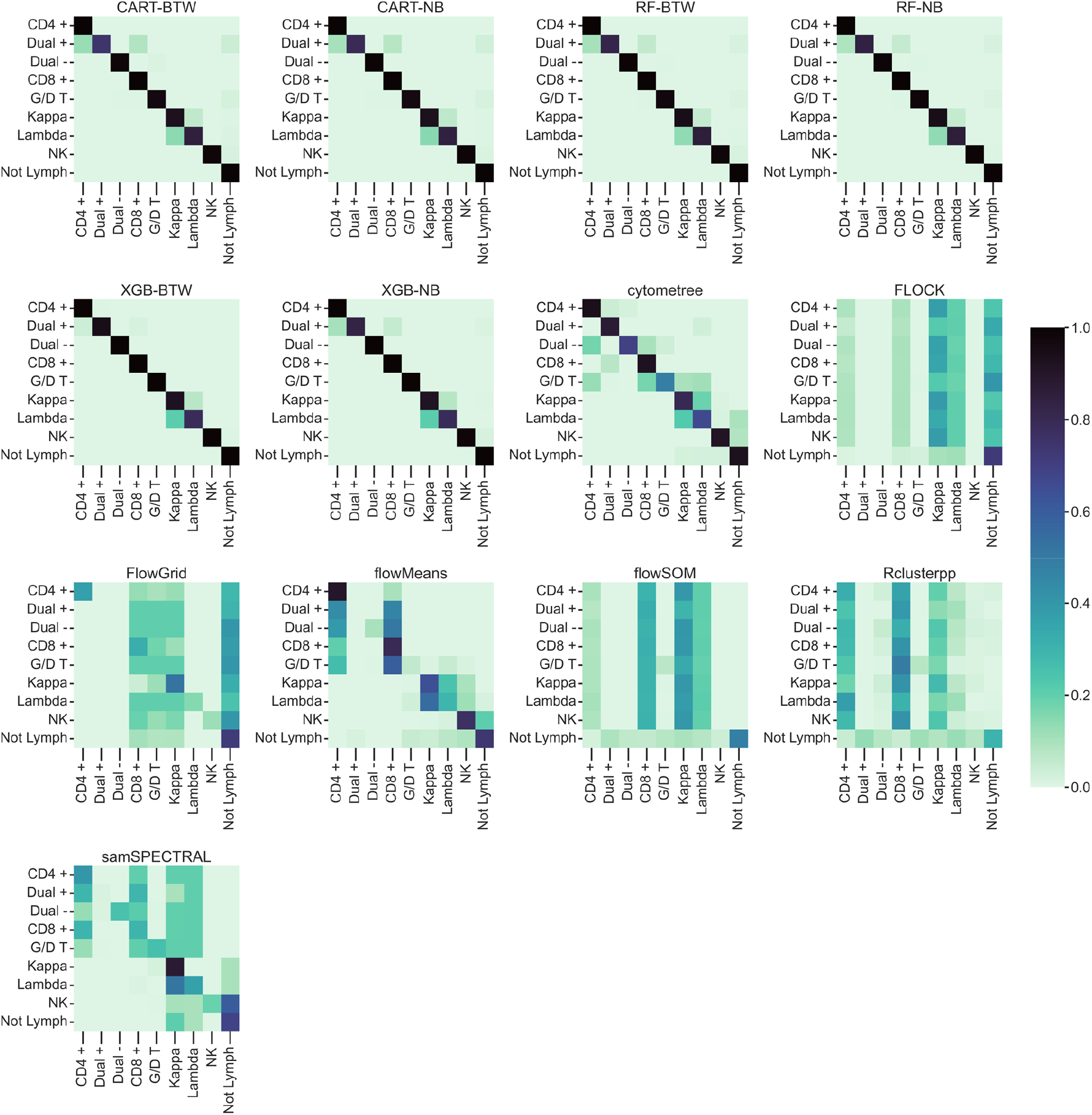
Mean aggregated confusion matrices for all classification algorithms and clustering methods. X-axis shows the predicted labels, y-axis shows the true labels for each class. Colour indicates proportion of predicted labels normalised by the number of true labels per class. CART, Random Forest (RF) and XGBoost (XGB) were trained once with models weighting classes inversely proportional to their frequencies within the dataset during model training (Balanced Training Weights = BTW), and once without (No Balancing = NB). Classification algorithms were cross validated over 10 folds with a labelled dataset containing events from 152 samples. Clustering methods were run on 10 randomly chosen samples from the labelled dataset, identified clusters were matched to labelled populations using the Hungarian assignment algorithm.

“Not lymphocyte” showed the highest mean F1 score across all methods and algorithms (0.82 ± 0.09). Other than “CD4/CD8 + T cell”, only two classes showed mean F1 scores of less than 0.98 for the classification algorithms, these were “Kappa B cell” and “Lambda B cell” which had mean F1 scores of 0.88 (± 0.07) and 0.85 (± 0.11), respectively. Confusion matrices for classification algorithms show that the B cell classes were frequently mislabelled as one another, with areas of darker green around the B cell classes. Figure 6 shows the kernel density estimation (KDE) of the B cell subtype distributions. KDE can be used to estimate the distribution of data and shows the overlap of Kappa and Lambda B cell distributions where these values are very similar.

**Fig 6.**
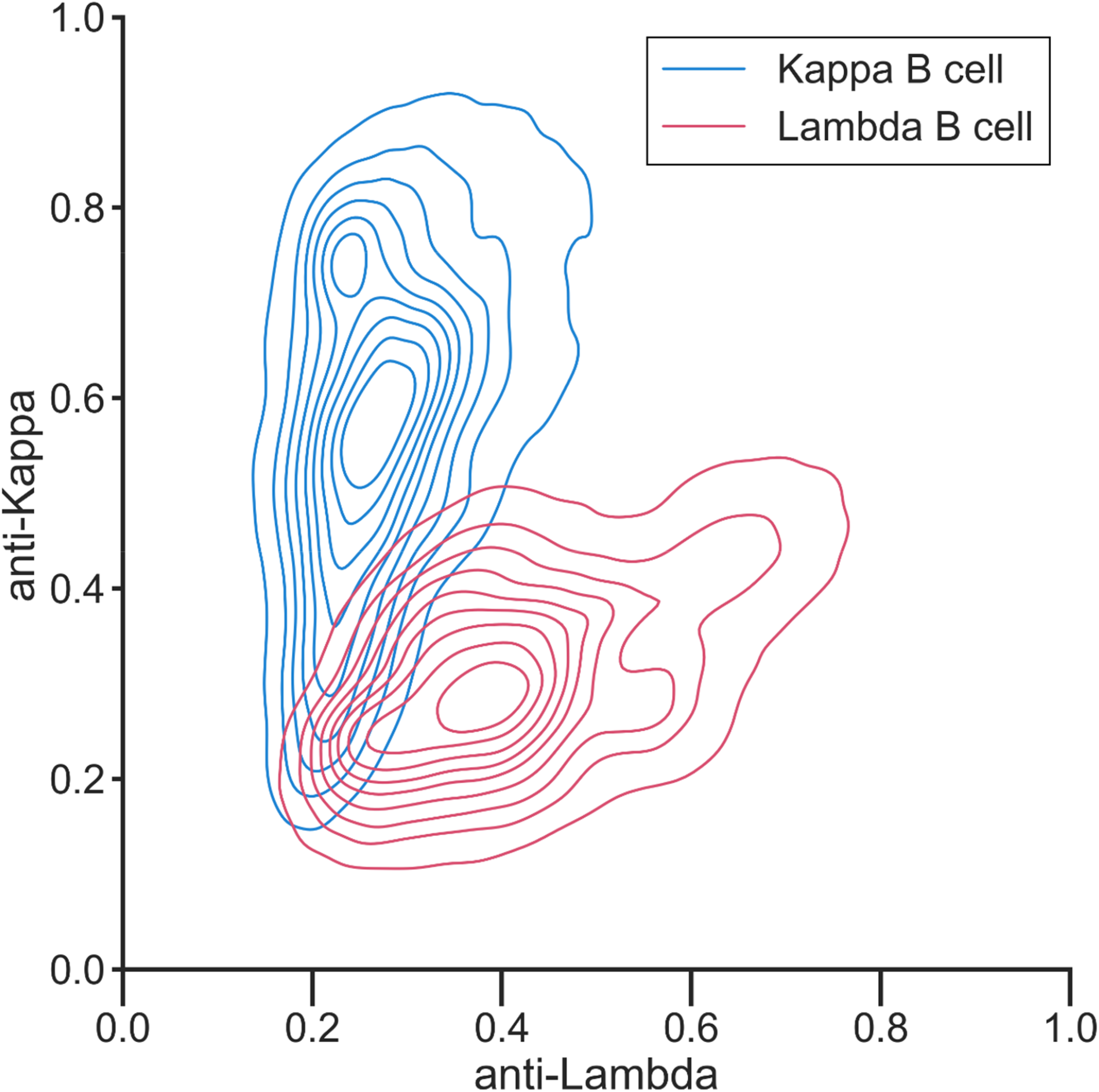
Kernel density estimation of B cell subtypes in dataset. Kernel density estimation of a randomly sampled 5% of the dataset showing the overlap between Kappa and Lambda B cell populations. B cells events are labelled as either Kappa or Lambda subtype depending on marker expression of their most proximal subpopulation, despite individual events of both subtypes frequently expressing identical phenotypic signatures.

Example predictions for a sample are plotted in Fig 7, demonstrating CART’s effectiveness at replicating manual analysis. A highlighted region show how cytometree has failed to cluster groups of events which have been assigned to T cell subtype labels by the Hungarian assignment algorithm, in comparison to the labelled data. The second highlighted region shows how flowMeans has clustered events together which contain a mixture of lymphocytes and other cell types, though these have been assigned to various lymphocyte subtypes by the Hungarian assignment algorithm. Though cytometree showed the highest f1 score of any of the clustering methods, it still resulted in misclassifications which could negatively impact diagnosis, suggesting that an F1 score of 0.6 is still not accurate enough to be clinically useful.

**Fig 7.**
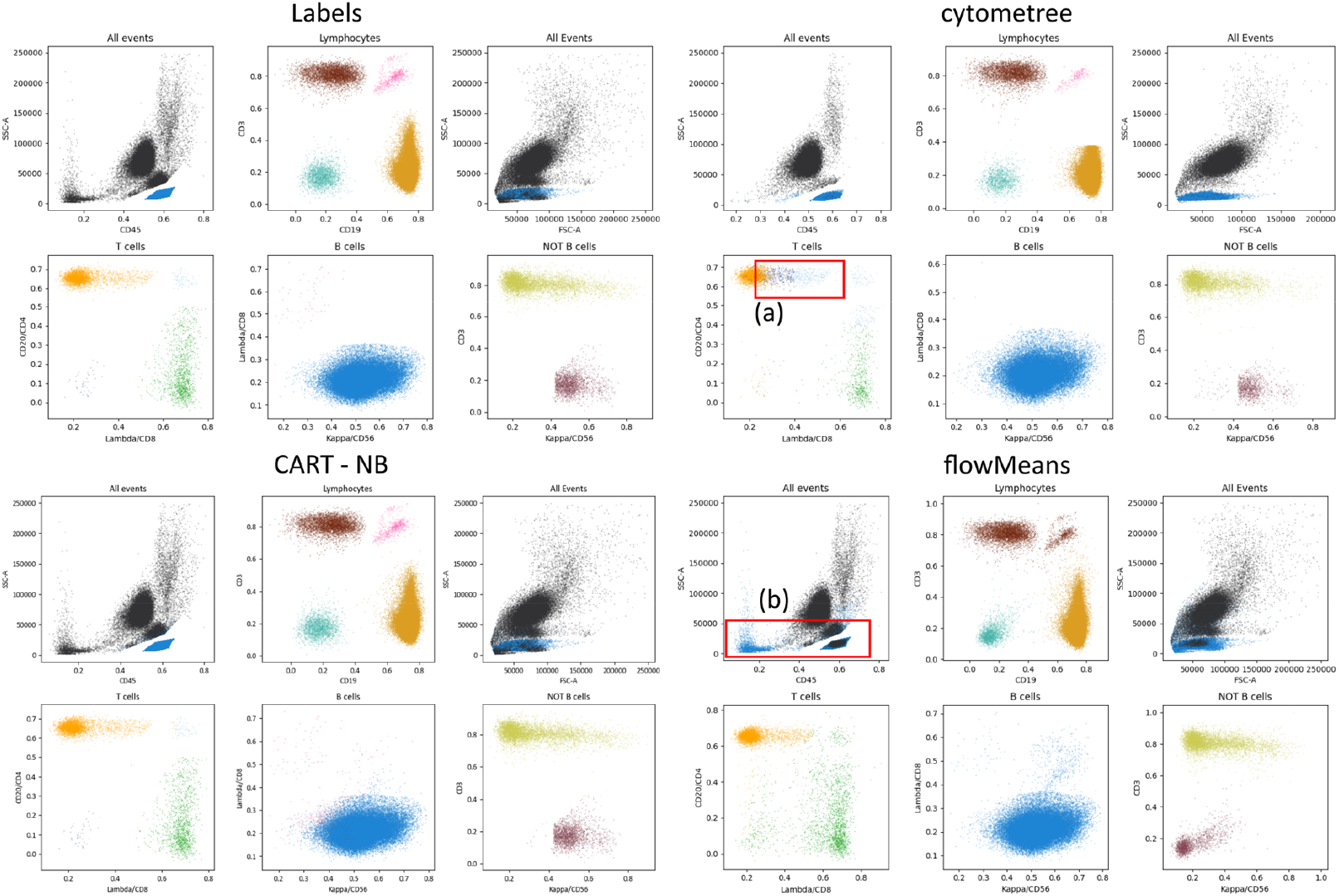
Labels and example output from CART and the best performing clustering methods. Top left, plotted labels from an example sample in the labelled dataset. Clockwise from top right, plotted predictions of cytometree, flowMeans and CART (trained with all instances being balanced equally i.e. No Balancing). Each plot shows the gating strategy for the Lymphoid Screening Tube panel which was used for all samples in the study, each colour indicates a different label given to each event. Initially, lymphocytes are identified from all events, before being gated into B, T and NK cell subtypes. CART was trained on 137 other samples. Clusters identified by clustering methods were matched to labelled populations using the Hungarian assignment algorithm. (a) Highlights cytometree incorrectly labelling subtypes of T cell. (b) Highlights flowMeans incorrectly labelling events which are not lymphocytes as lymphocytes.

## Discussion

The purpose of this research was to develop an automated event labelling approach for FC analysis which utilises tree-based classification algorithms, and test against previously published clustering methods.

### Tree-based classification algorithms perform well at automated FC analysis

The classification algorithms showed higher mean F1 scores than clustering methods, though the difference was insignificant between classification algorithms. Classification algorithm prediction time was comparable to clustering methods, though training time was variable between classification algorithms, with CART models taking, on average, less than 7.5 minutes to train on data generated from 137 samples, whereas XGB models took, on average, over 28 hours on FC data generated from 110 samples.

CART and RF both showed high mean F1 scores of over 0.94, short mean prediction times of less than 25 seconds, and short mean training times of less than 2 hours. CART, in particular, showed the shortest prediction and training time, therefore combined with very high F1 scores it was perhaps the best all-round tree-based algorithm of those evaluated. CART achieved similar F1 scores to RF and XGB, suggesting the task was not complex enough to justify the use of the more complex RF and XGB models. XGB suffered from long training times, likely due to the process of randomised grid-search to optimise hyper-parameters (HPO). HPO is the practice of optimising certain model parameters to improve performance. Randomised grid search is an HPO method involving repeatedly evaluating the model using various combinations of hyper-parameters with the aim to identify those which are optimal. Decreasing the hyper-parameter search space or performing HPO on a smaller subset of data would likely decrease training time, though could result in poorer model performance.

Models weighting classes inversely proportional to their frequencies during training provided no improvement in F1 score performance. XGB models trained with weighted classes exhibited higher mean recall of all classes, though particularly “CD4/CD8 + T cell” which was the lowest frequency class in the dataset, making up only 0.03% overall. Conversely the mean precision of these models was lower in all classes, and particularly “CD4/CD8 + T cell” (Supplementary Table 1 and 2), suggesting that minority classes were more likely to be predicted by the XGB weighted model, compared to the unweighted, regardless of whether the prediction was correct. A weighted XGB model may be preferred to other algorithms if higher recall was valued over precision or F1 score, for example if it was deemed critical that the presence of a minority certain cell type was identified, and therefore false positives were preferable to false negatives.

### Clustering methods showed poor F1 score at automated analysis of FC data generated from samples with suspected haematological malignancy

Of clustering methods evaluated, only cytometree achieved a mean F1 score of above 0.4. FLOCK, flowMeans, flowSOM, Rclusterpp and samSPECTRAL were all selected for evaluation due to performing well in previous comparison studies where some of the methods achieved mean F1 scores of over 0.9 [25]. The discrepancy between the high F1 scores previously reported and the low F1 scores reported by this paper could be explained by the differing datasets. The method of gating the panel used this study, the Lymphoid Screening Tube (LST) (described in Supplementary Information), involves immediately gating out events that are not deemed to be lymphocytes based on SSC-A and CD45. These events were labelled “Not lymphocyte” and made up 61% of the dataset. They consist of a variety of different cell types including granulocytes and monocytes, as well as debris. The evaluated clustering methods have no capacity to understand the events which are and are not relevant to an FC analyst. Therefore, they may be returning valid cell populations which are not of interest, for example, in the case of the LST panel, anything which is not a lymphocyte population. This effect, combined with the use of the Hungarian assignment algorithm, likely explains why “Not lymphocyte” was the class which exhibited the highest mean F1 score by clustering methods. A clustering method would be rewarded for returning the largest population of cells, as long as they were not lymphocytes, which 61% of the dataset was not, as the Hungarian assignment algorithm would then match this population as being “Not lymphocyte”.

Cytometree achieved the highest mean F1 score of any clustering method, while also exhibiting a mean prediction time of less than 20 seconds. This was of particular interest as it was the only tree-based clustering method evaluated. Gating is analogous to the decision-making process tree-based algorithms use, and the good performance of all tree-based methods involved in the study suggests FC automated gating is a task well suited to this class of algorithms.

### Tree-based models performed less well on Kappa and Lambda B cells

“CD4/CD8 + T cell” showed the lowest F1 score of any individual class across all algorithms, though made up the lowest frequency of all classes in the dataset. The only other two classes with mean F1 scores below 0.98 for classification algorithms were “Kappa B cell” and “Lambda B cell”, which had mean F1 scores of 0.88 (± 0.07) and 0.85 (± 0.11), respectively. However, these made-up large proportions of the dataset (*“Kappa B cell”:* 12.29% and *“Lambda B cell”:* 11.67%), suggesting lack of training data was not the factor limiting performance. Furthermore “Kappa B cell” and “Lambda B cell” were the classes which cytometree most frequently confused. Due to the nature of manual gating, events are not considered in isolation. Instead, several thousand events at minimum, are plotted and gated simultaneously. In contrast, in this study, the classification algorithms were trained and made predictions on a single event at a time, not considering other events and populations. The gating procedure is demonstrated in the Supplementary Information. Kappa and Lambda B cells often exhibit similar phenotypic signatures when stained with the LST panel and may be even more similar in certain conditions, such as CLL where Kappa or Lambda expression are known to be weak [33]. B cell events are manually gated by considering the position of the centroid of the sub-population which the events are closest in proximity to i.e. an event classified as a “Kappa B cell” or a “Lambda B cell” may have similar values, but if the event is closer to a cluster of cells with a centroid located more towards Kappa +, then the event may be labelled “Kappa B cell” and vice versa for “Lambda B cell”. Therefore, as the classification algorithms learn and make predictions based solely on the values of a single event, they do not possess the same information which led to labelling the events.

### Future Research

Tree-based models may show the required accuracy and prediction times to be suitable for automated FC for the diagnosis of haematological malignancy. Additionally, classification algorithms do not suffer from the downside of having to further label clustered sub-populations, saving further processing time. However, there are challenges to overcome. The classification algorithms trained during this study are only able to make predictions on the LST panel, predicting only the classes that the data was trained on. Labelling the data required for training is time consuming.

Therefore, future research should focus on testing tree-based classifiers on datasets generated by other panels. In particular, due to the variability associated with manual gating a “consensus dataset” made up of samples gated by multiple operators to act as “gold standard” labels would be preferable to a single operator labelling the data, as was the case during the present study [8].

When considering the poor performance of the other clustering methods, cytometree performed well and may provide a platform for further development towards a goal of accurate, fast, fully automated FC analysis which would also not require labelled data as classification algorithms do.

Tree-based methods, both cytometree and classification algorithms, were the best performing methods, though struggled with B cell classes more than other classes. B cell classes are gated with particular consideration towards the surrounding cell sub-populations. Therefore, future research may also look to consider how tree-based methods could be combined with density or mixture model-based approaches to pair the accuracy of tree-based algorithms while considering the surrounding cell sub-populations.

In conclusion, tree-based classification algorithms could provide a solution for automated FC analysis of known populations. When used for the diagnosis of haematological malignancies, this could reduce diagnostic bottlenecks [13]. CART especially, showed short prediction and training times, and very high F1 scores, comparable with RF and XGB. The only tree-based clustering method evaluated, cytometree, was the best performing of the clustering methods, suggesting that tree-based methods are well suited to FC analysis. Future research should prioritise further evaluation of the discussed methods on other FC datasets, as well as looking to improve the performance of tree-based classification methods by providing information of on surrounding sub-populations, rather than solely single events.

## Methods

### Ethics Statement

This study was approved by the University of Liverpool for Sponsorship in Dec 2020 (UoL001599) and Integrated Research Application System (IRAS) approval was granted (project ID: 290362).

### Dataset

The Haemato-Oncology Diagnostics Service (HODS) based at the Royal Liverpool University Hospital carries out primary reporting of haematological malignancies for the two million people within the Merseyside and Cheshire area. One hundred and fifty-two fcs 2.0 files were used for the dataset which had been collected by HODS between the dates of February 2018 and March 2020. The data was generated from peripheral blood samples from patients with either an abnormal full blood count result or suspected haematological malignancies for whom FC had been performed as part of the diagnostic pathway. Samples had been stained with the EuroFlow LST panel (BD Biosciences, San Jose, CA) for the purpose of identifying aberrant B, T and NK cell lineages [28]. All samples were analysed on FACSCanto cytometers (BD Biosciences, San Jose, CA). Each file had been compensated and contained 100,000 events from a single patient sample. As part of HODS’s standard operating procedure, each file had undergone a quality control process within HODS which included samples being checked for any signs of degradation during histopathological examination and the ability to observe distinguishable clusters on SSC and CD45 parameters of FC data.

### Gating

Each fcs file was gated by a single trained FC operator using manual gating software (FlowJo, BD Biosciences, San Jose, CA). Each sample was gated to label events the operator was confident of the identity of. One of nine labels was given to these events: “Not lymphocyte”, “Kappa B cell”, “Lambda B cell”, “CD4+ T cell”, “CD8+ T cell”, “CD4/CD8 + T cell”, “CD4/CD8 – T cell”, “Gamma delta T cell”, “CD56+ NK cell”. Details of gating strategy are included in Supplementary Information. Each class of labelled event from a file was exported to a csv.

### Pre-processing

The labelled events from each sample were combined and fluorescence channels were transformed using the Logicle transformation, while FSC and SSC remained linear [34]. The Logicle transformation has been suggested as the preferred transformation for FC data, resulting in fewer misclassified events than other popular transformations [35]. The transformed data from each sample was plotted in 2D scatter plots and checked for visual similarity to the equivalent unlabelled sample to ensure that transformation was successful. No pre-gating was performed, all ungated labelled events were used for evaluation. Table 2 summarises the labelled events in the dataset.

**Table 2.**
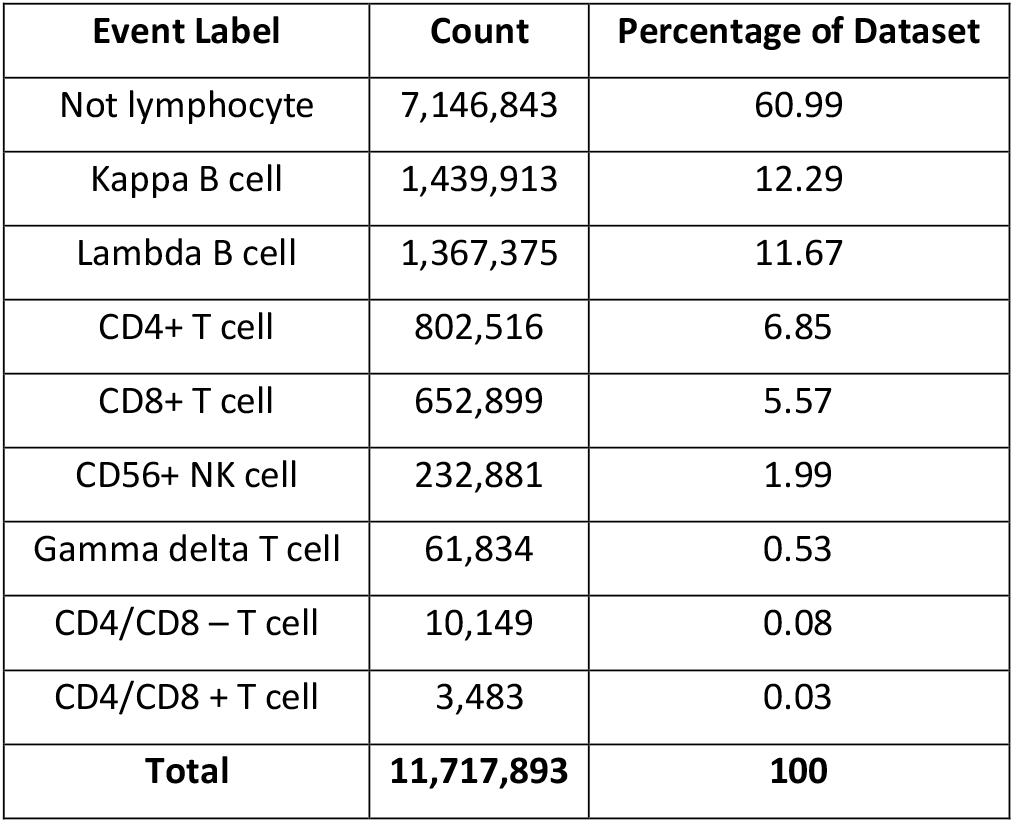
The frequency of each class label within the dataset.

### Classification algorithms and training

Ten-fold cross-validation was used to measure the performance of the models. Folds and splits were made between samples, rather than events, meaning events within a sample would not appear in both train and test sets within a fold. This was done to simulate how an algorithm would be trained in practice i.e., the data from an entire sample would be analysed together. For CART and RF, there was a 90/10 train-test split in each fold. For XGB, there was a 72/18/10 train-test-validation split in each fold, as 10% was used for validation, and of the resulting 90%, 80% was used for training and the rest for testing.

As sub-populations identified by FC differ in size, classes were heavily imbalanced. Tree based methods tend to work best with balanced data [36]. Therefore, analysis was repeated twice, once with equal weighting given to all instances during model training, and once with models weighting classes inversely proportional to their frequencies during model training. Hyperparameters of XGB were tuned using randomised grid-search prior to model training. All analysis was performed in Python (version 3.9) [37]. The sklearn package (0.24.2) was used for CART and RF, and xgboost package (1.5.1) for XGB [31,38].

### Clustering methods

To identify previously published automated FC analysis methods, PubMed was searched with the search term “(“flow cytometry” OR “FCM”) AND (“automat*” OR “clustering” OR “classification” OR “machine learning” OR “random forest” OR “CART” OR “decision tree*”)”. Search results were assessed for relevance and relevant references were explored. The following criteria was used for selecting a short list of clustering methods for benchmarking:

1. The method was designed to be used with FC data to identify or cluster similar events so they can be labelled.
2. The method was freely available to use either as source code or application.
3. The method was straightforward to use, meaning; either default or suggested parameters were detailed, and the method ran either without error, or with errors which could be fixed without modification to source code.
4. Either the method had previously been independently assessed or had been released since the most recently published critical assessment paper in 2016 [25]. I.e., there had been no opportunity for it to be independently assessed.

All methods which had previously been independently assessed were included in either one of two papers [8,25]. Due to the large number of methods which had previously been assessed, only the three best performing methods based on mean F1 score from both papers were included for benchmarking. This resulted in a shortlist of nine clustering methods selected for benchmarking, these methods used a variety of approaches (Table 3). BayesFlow and X-shift could not be successfully installed for testing, therefore seven methods were benchmarked.

**Table 3.** Overview of clustering methods selected for benchmarking evaluation.

| Method | Description | Availability | Rationale for inclusion | Ref |
| --- | --- | --- | --- | --- |
| BayesFlow | Bayesian hierarchical modelling and Gaussian Mixture Modelling | Python library from GitHub | Published since the most recent critical assessment | [39] |
| cytometree | Binary trees where nodes are sub-populations of cells | R package | Published since the most recent critical assessment | [40] |
| FLOCK | Density based clustering, followed by merging | C source code | 3<sup>rd</sup> best performing method during Aghaeepour *et al.* (2013) | [41] |
| FlowGrid | Density based clustering, followed by merging | Python library from GitHub | Published since the most recent critical assessment | [42] |
| flowMeans | K-means clustering, followed by merging | R package | Best performing method during Aghaeepour *et al.* (2013) | [43] |
| flowSOM | Self-organising map, followed by merging using hierarchical clustering | R package | 2<sup>nd</sup> best performing method during Weber *et al.* (2016) | [44] |
| Rclusterpp | Hierarchical clustering | R package | 3<sup>rd</sup> best performing method during Weber *et al.* (2016) | [45] |
| samSPECTRAL | Speed optimised spectral clustering | R package | 2<sup>nd</sup> best performing method during Aghaeepour *et al.* (2013) | [46] |
| X-shift | K – nearest – neighbours based | Standalone application run through CLI | Best performing method during Weber *et al.* (2016) | [47] |

Ten samples were randomly selected from the labelled dataset to evaluate clustering methods performance on. Not all 152 files in the dataset were used as some of the clustering methods can take multiple hours to run [25]. Each method was run five times on each sample due to the stochastic nature of some of the methods. Some methods allowed the number of output clusters which were to be identified by the method to be specified, before running the method. If allowed, this was set to 9, matching the number of classes in the dataset. Clusters identified by the methods were matched to the labelled populations using the Hungarian assignment algorithm which maximises the sum of F1 scores across populations, allowing no population to be matched more than once [25].

### Evaluation metrics

Precision is the proportion of instances of a class which are identified correctly, and recall is the proportion of actual class samples which are identified correctly. F1 score is the harmonic mean of precision and recall. In this paper, macro averaged F1 score was calculated (averaged over the set of all classes) and is a preferred evaluation metric when classes are imbalanced [48]. The training time, in seconds, of each classifier was recorded. Measurements for running time were made on a cluster with a cumulative 50 cores and 100GB of RAM.

### Statistical Analysis

A two-way ANOVA was performed to analyse the effect of classifier algorithm and weighting training instances on F1 score. Statistical significance was set as p < 0.05. Data in figures, tables and text are presented as means ± standard deviation.

## Data Availability

Code for running experiments is available on GitHub (https://github.com/alex-rothwell/classification_evaluation_flow_cytometry). Patient data is unavailable due to ethical restrictions imposed by IRAS.

## Acknowledgments

We acknowledge the Liverpool Clinical Labs’ Haemato-Oncology Diagnostic Service for providing the FCS files used for the dataset.

## Data Availability

Code for running experiments is available on GitHub at (https://github.com/alex-rothwell/classification_evaluation_flow_cytometry). Patient data is unavailable due to ethical restrictions imposed by IRAS (project ID: 290362).

## Supplementary Information

### Gating Strategy

1. From all events within an fcs file, lymphocytes were identified as being CD45+ / SSC low. All other events were labelled “Not lymphocytes”.

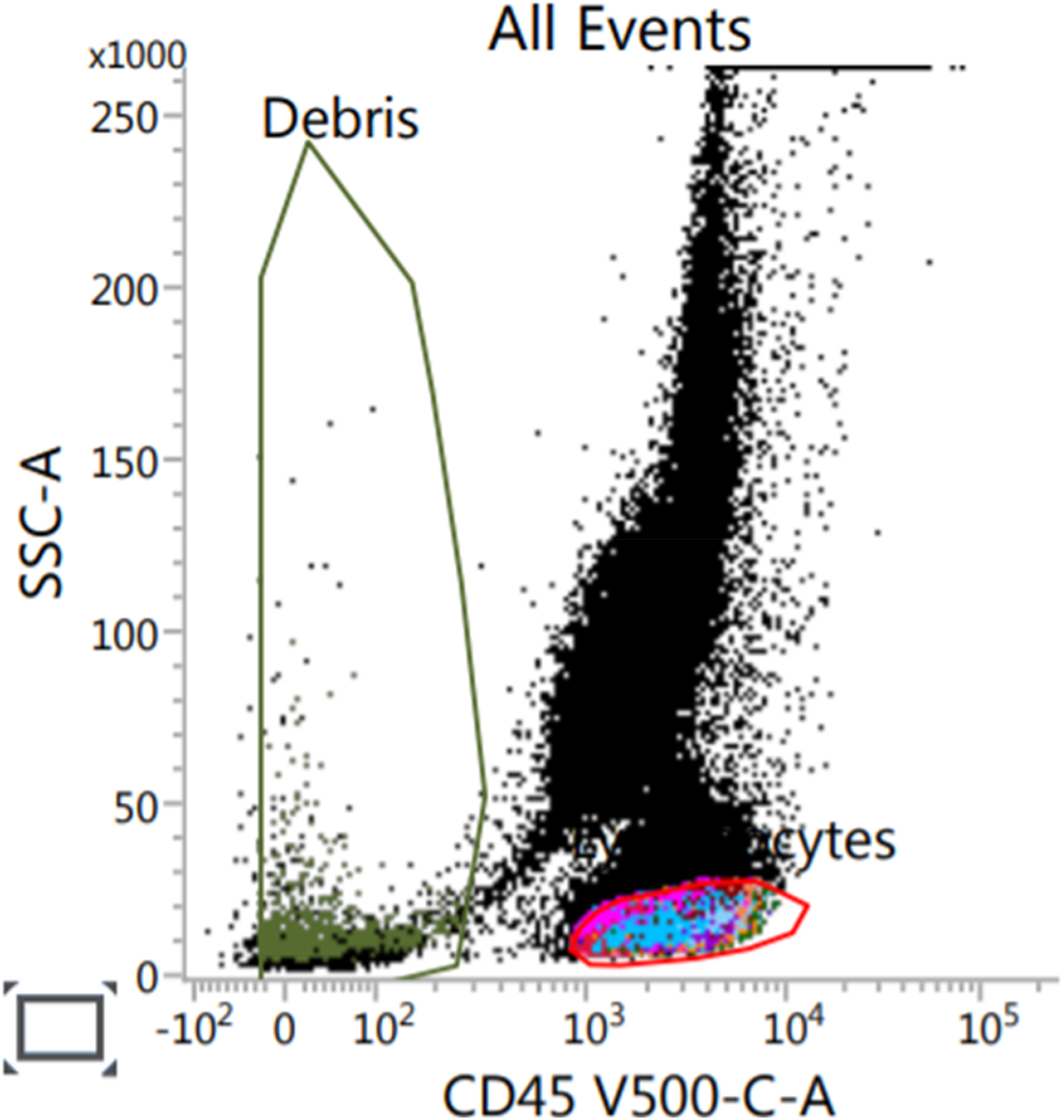
2. From lymphocytes, T cells were identified as being CD3+ / CD19-, Gamma Delta T cells as being CD3+ / CD19+, and B cells as being CD3- / CD 19+.

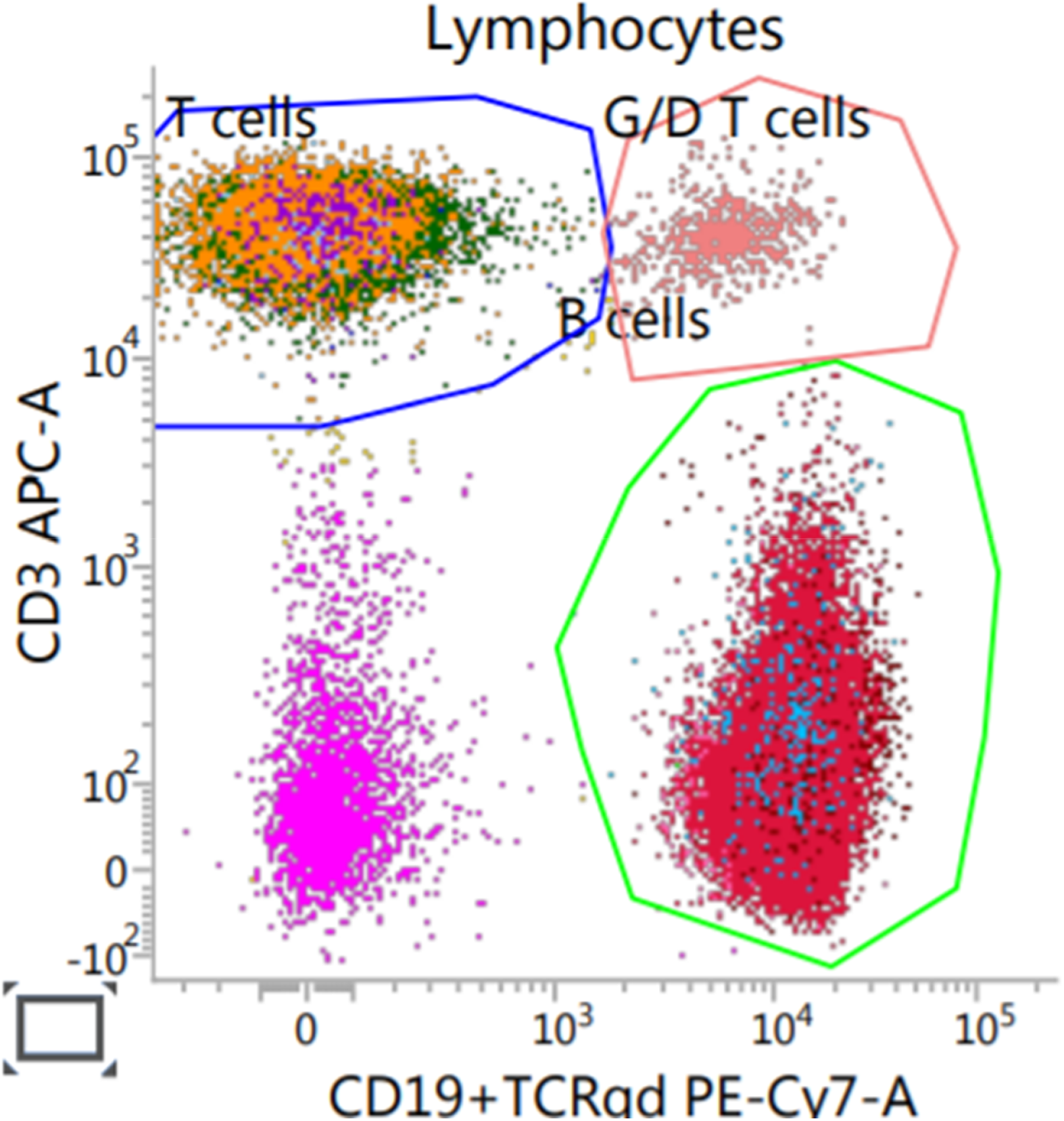
3. From T cells, CD4+ T cells were identified as being CD4+ / CD8-, CD8+ T cells as being CD4-/ CD8+, CD4/CD8-T cells and CD4/CD8 + T cells were identified as described.

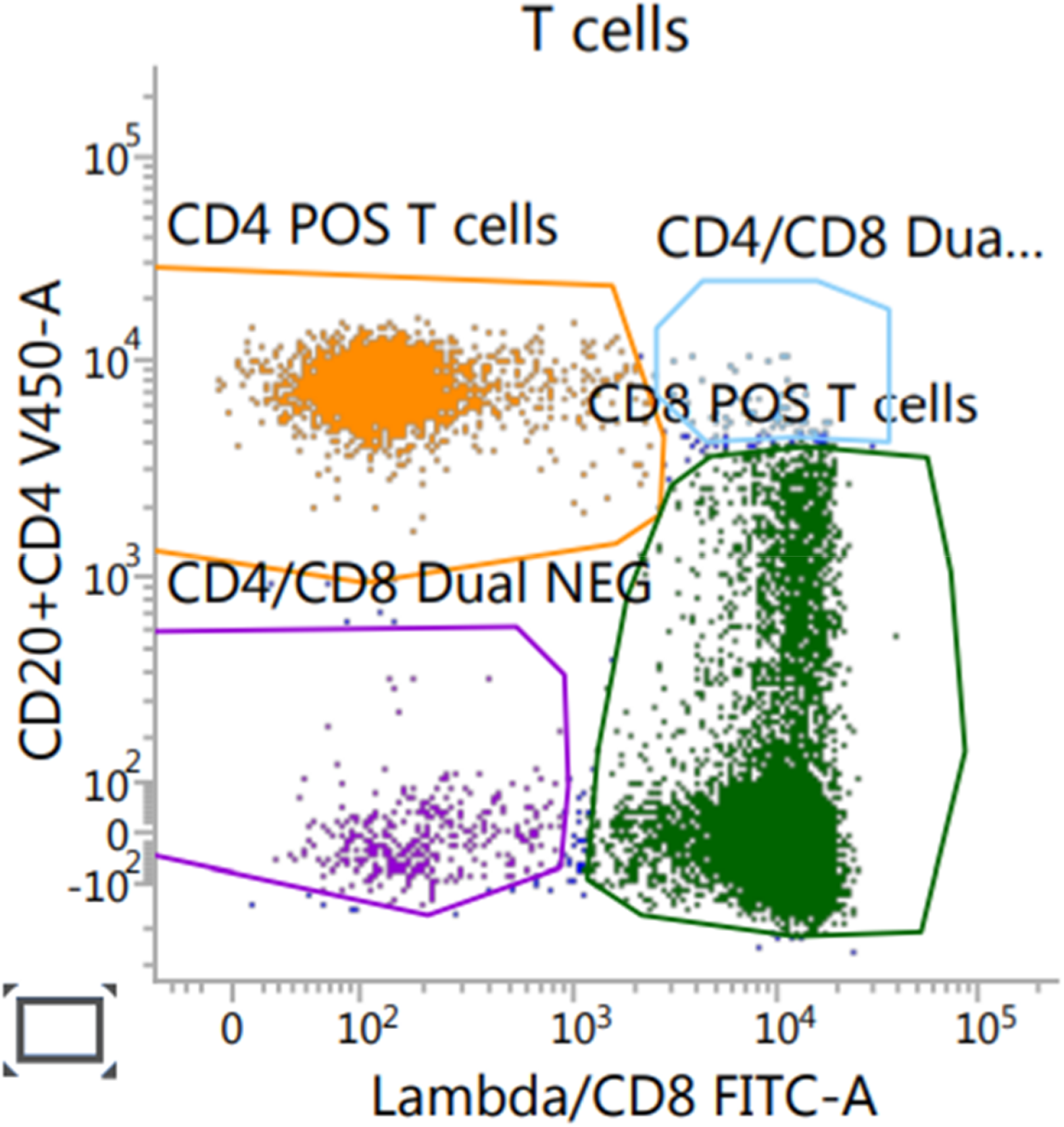
4. From B cells, Kappa B cells being identified as part of a Kappa+ population, Lambda B cells as being part of a Lambda + population.

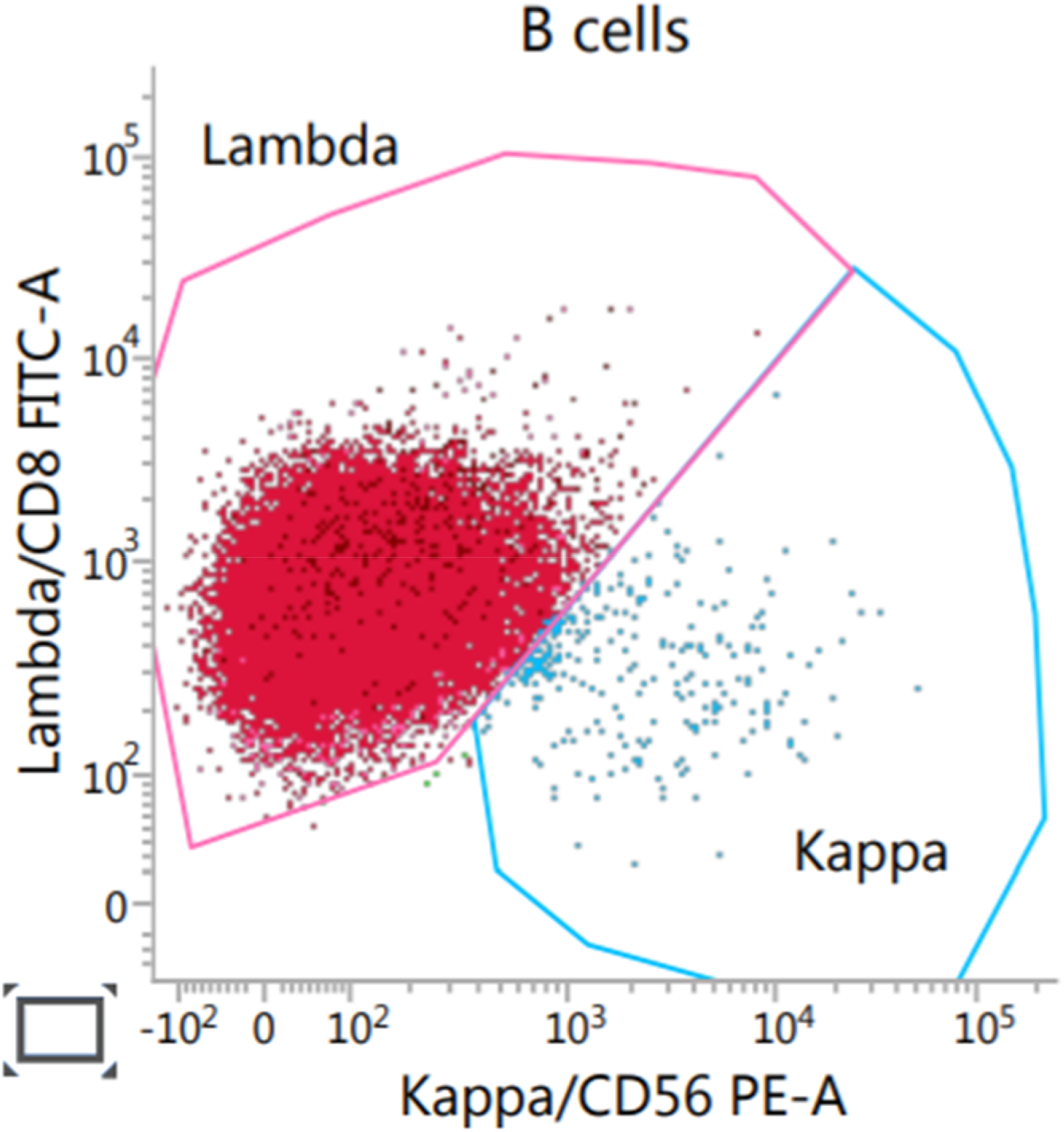
5. From lymphocytes, which had not been labelled as B cells, NK cells were identified as CD56+.

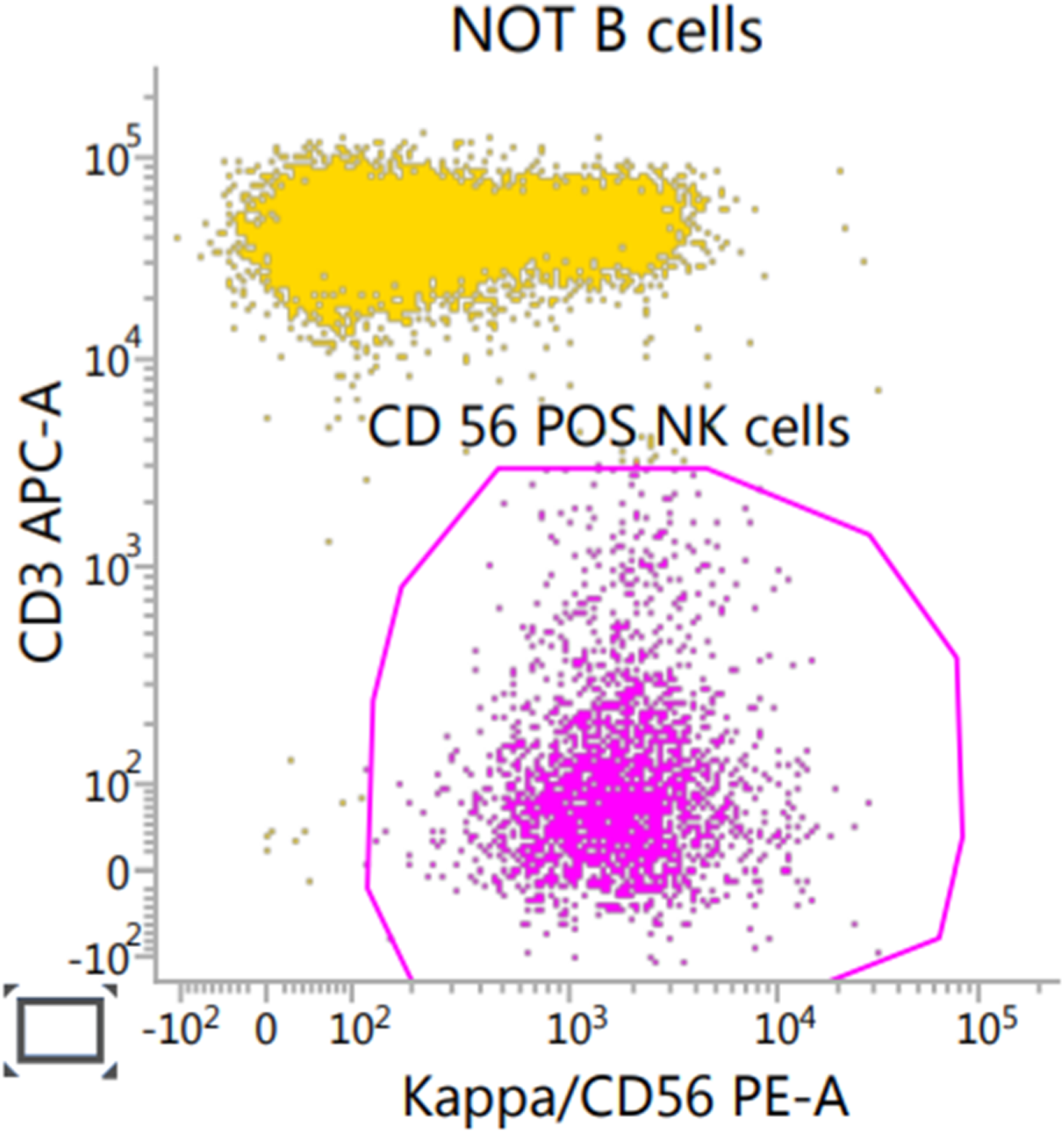

---

**Supplementary Table 1.** Mean (± SD) recall for each class for each classification algorithm and clustering method.

| Method \ Class | CD4 + T cell | Dual + T cell | Dual – T cell | CD8 + T cell | G/D T cell | Kappa B cell | Lambda B cell | NK cell | Not Lymph | Overall |
| --- | --- | --- | --- | --- | --- | --- | --- | --- | --- | --- |
| Random Forest-No Balancing | 1.0 (± 0.0) | 0.84 (± 0.15) | 0.99 (± 0.01) | 1.0 (± 0.0) | 0.98 (± 0.02) | 0.95 (± 0.07) | 0.86 (± 0.2) | 0.99 (± 0.01) | 1.0 (± 0.0) | **0.96 (± 0.05)** |
| Random Forest-Balanced Training Weights | 1.0 (± 0.0) | 0.82 (± 0.16) | 0.99 (± 0.01) | 1.0 (± 0.0) | 0.98 (± 0.03) | 0.95 (± 0.07) | 0.86 (± 0.2) | 0.99 (± 0.02) | 1.0 (± 0.0) | **0.95 (± 0.06)** |
| XGBoost-No Balancing | 1.0 (± 0.0) | 0.84 (± 0.1) | 1.0 (± 0.0) | 1.0 (± 0.0) | 0.99 (± 0.0) | 0.92 (± 0.06) | 0.79 (± 0.09) | 0.98 (± 0.01) | 1.0 (± 0.0) | **0.95 (± 0.03)** |
| CART-Balanced Training Weights | 1.0 (± 0.0) | 0.75 (± 0.15) | 0.99 (± 0.02) | 1.0 (± 0.0) | 0.97 (± 0.03) | 0.93 (± 0.07) | 0.85 (± 0.18) | 0.98 (± 0.02) | 1.0 (± 0.0) | **0.94 (± 0.05)** |
| CART-No Balancing | 1.0 (± 0.0) | 0.8 (± 0.13) | 0.99 (± 0.0) | 1.0 (± 0.0) | 0.96 (± 0.03) | 0.93 (± 0.07) | 0.85 (± 0.18) | 0.99 (± 0.01) | 1.0 (± 0.0) | **0.95 (± 0.05)** |
| XGBoost-Balanced Training Weights | 1.0 (± 0.0) | 0.95 (± 0.08) | 1.0 (± 0.0) | 1.0 (± 0.0) | 1.0 (± 0.0) | 0.93 (± 0.05) | 0.79 (± 0.08) | 0.99 (± 0.0) | 1.0 (± 0.0) | **0.96 (± 0.03)** |
| cytometree | 0.87 (± 0.09) | 0.55 (± 0.43) | 0.58 (± 0.36) | 0.6 (± 0.14) | 0.19 (± 0.13) | 0.62 (± 0.33) | 0.56 (± 0.35) | 0.71 (± 0.18) | 0.65 (± 0.16) | **0.59 (± 0.24)** |
| flowMeans | 0.9 (± 0.3) | 0.0 (± 0.0) | 0.1 (± 0.3) | 0.8 (± 0.4) | 0.06 (± 0.13) | 0.64 (± 0.45) | 0.29 (± 0.45) | 0.77 (± 0.38) | 0.73 (± 0.13) | **0.48 (± 0.28)** |
| samSPECTRAL | 0.4 (± 0.49) | 0.01 (± 0.03) | 0.25 (± 0.37) | 0.3 (± 0.46) | 0.26 (± 0.39) | 0.88 (± 0.3) | 0.37 (± 0.46) | 0.19 (± 0.38) | 0.66 (± 0.44) | **0.37 (± 0.37)** |
| FlowGrid | 0.32 (± 0.1) | 0.0 (± 0.0) | 0.0 (± 0.0) | 0.3 (± 0.36) | 0.2 (± 0.4) | 0.51 (± 0.42) | 0.14 (± 0.29) | 0.12 (± 0.14) | 0.55 (± 0.16) | **0.24 (± 0.21)** |
| flowSOM | 0.1 (± 0.29) | 0.0 (± 0.0) | 0.0 (± 0.0) | 0.31 (± 0.45) | 0.07 (± 0.18) | 0.4 (± 0.49) | 0.21 (± 0.4) | 0.0 (± 0.0) | 0.46 (± 0.15) | **0.17 (± 0.22)** |
| FLOCK | 0.09 (± 0.28) | 0.0 (± 0.0) | 0.0 (± 0.0) | 0.1 (± 0.29) | 0.01 (± 0.02) | 0.38 (± 0.47) | 0.2 (± 0.4) | 0.0 (± 0.0) | 0.72 (± 0.28) | **0.17 (± 0.19)** |
| Rclusterpp | 0.32 (± 0.28) | 0.0 (± 0.0) | 0.05 (± 0.13) | 0.41 (± 0.37) | 0.08 (± 0.15) | 0.28 (± 0.32) | 0.12 (± 0.24) | 0.02 (± 0.03) | 0.29 (± 0.05) | **0.17 (± 0.17)** |
| **Mean: Clustering method** | **0.43 (± 0.26)** | **0.08 (± 0.07)** | **0.14 (± 0.17)** | **0.4 (± 0.35)** | **0.12 (± 0.2)** | **0.53 (± 0.4)** | **0.27 (± 0.37)** | **0.26 (± 0.16)** | **0.58 (± 0.2)** | **0.31 (± 0.24)** |
| **Mean: Classification algorithm** | **1.0 (± 0.0)** | **0.83 (± 0.13)** | **0.99 (± 0.01)** | **1.0 (± 0.0)** | **0.98 (± 0.02)** | **0.93 (± 0.06)** | **0.83 (± 0.16)** | **0.99 (± 0.01)** | **1.0 (± 0.0)** | **0.95 (± 0.04)** |
| **Mean: All** | **0.69 (± 0.14)** | **0.43 (± 0.1)** | **0.54 (± 0.09)** | **0.68 (± 0.19)** | **0.52 (± 0.12)** | **0.72 (± 0.24)** | **0.53 (± 0.27)** | **0.6 (± 0.09)** | **0.78 (± 0.11)** | **0.61 (± 0.15)** |

**Supplementary Table 2.**
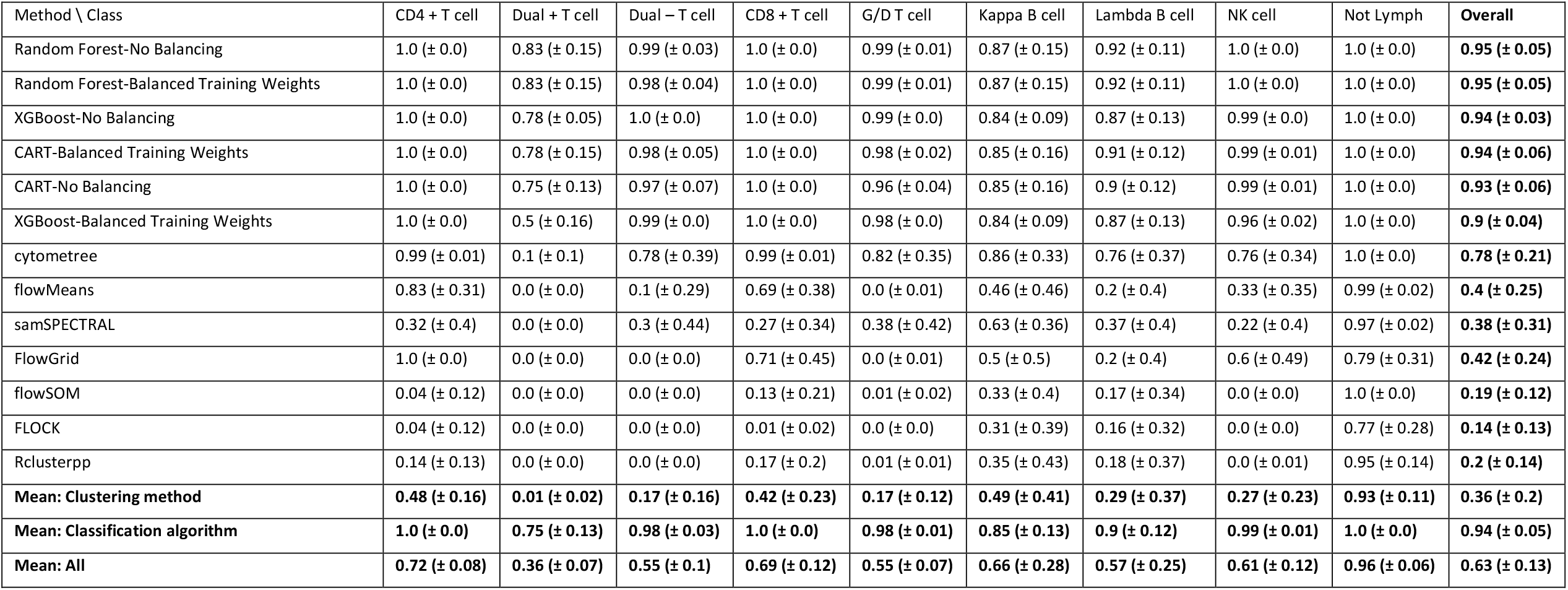
Mean (± SD) precision for each class for each classification algorithm and clustering method.

| Method \ Class | CD4 + T cell | Dual + T cell | Dual – T cell | CD8 + T cell | G/D T cell | Kappa B cell | Lambda B cell | NK cell | Not Lymph | Overall |
| --- | --- | --- | --- | --- | --- | --- | --- | --- | --- | --- |
| Random Forest-No Balancing | 1.0 (± 0.0) | 0.83 (± 0.15) | 0.99 (± 0.03) | 1.0 (± 0.0) | 0.99 (± 0.01) | 0.87 (± 0.15) | 0.92 (± 0.11) | 1.0 (± 0.0) | 1.0 (± 0.0) | **0.95 (± 0.05)** |
| Random Forest-Balanced Training Weights | 1.0 (± 0.0) | 0.83 (± 0.15) | 0.98 (± 0.04) | 1.0 (± 0.0) | 0.99 (± 0.01) | 0.87 (± 0.15) | 0.92 (± 0.11) | 1.0 (± 0.0) | 1.0 (± 0.0) | **0.95 (± 0.05)** |
| XGBoost-No Balancing | 1.0 (± 0.0) | 0.78 (± 0.05) | 1.0 (± 0.0) | 1.0 (± 0.0) | 0.99 (± 0.0) | 0.84 (± 0.09) | 0.87 (± 0.13) | 0.99 (± 0.0) | 1.0 (± 0.0) | **0.94 (± 0.03)** |
| CART-Balanced Training Weights | 1.0 (± 0.0) | 0.78 (± 0.15) | 0.98 (± 0.05) | 1.0 (± 0.0) | 0.98 (± 0.02) | 0.85 (± 0.16) | 0.91 (± 0.12) | 0.99 (± 0.01) | 1.0 (± 0.0) | **0.94 (± 0.06)** |
| CART-No Balancing | 1.0 (± 0.0) | 0.75 (± 0.13) | 0.97 (± 0.07) | 1.0 (± 0.0) | 0.96 (± 0.04) | 0.85 (± 0.16) | 0.9 (± 0.12) | 0.99 (± 0.01) | 1.0 (± 0.0) | **0.93 (± 0.06)** |
| XGBoost-Balanced Training Weights | 1.0 (± 0.0) | 0.5 (± 0.16) | 0.99 (± 0.0) | 1.0 (± 0.0) | 0.98 (± 0.0) | 0.84 (± 0.09) | 0.87 (± 0.13) | 0.96 (± 0.02) | 1.0 (± 0.0) | **0.9 (± 0.04)** |
| cytometree | 0.99 (± 0.01) | 0.1 (± 0.1) | 0.78 (± 0.39) | 0.99 (± 0.01) | 0.82 (± 0.35) | 0.86 (± 0.33) | 0.76 (± 0.37) | 0.76 (± 0.34) | 1.0 (± 0.0) | **0.78 (± 0.21)** |
| flowMeans | 0.83 (± 0.31) | 0.0 (± 0.0) | 0.1 (± 0.29) | 0.69 (± 0.38) | 0.0 (± 0.01) | 0.46 (± 0.46) | 0.2 (± 0.4) | 0.33 (± 0.35) | 0.99 (± 0.02) | **0.4 (± 0.25)** |
| samSPECTRAL | 0.32 (± 0.4) | 0.0 (± 0.0) | 0.3 (± 0.44) | 0.27 (± 0.34) | 0.38 (± 0.42) | 0.63 (± 0.36) | 0.37 (± 0.4) | 0.22 (± 0.4) | 0.97 (± 0.02) | **0.38 (± 0.31)** |
| FlowGrid | 1.0 (± 0.0) | 0.0 (± 0.0) | 0.0 (± 0.0) | 0.71 (± 0.45) | 0.0 (± 0.01) | 0.5 (± 0.5) | 0.2 (± 0.4) | 0.6 (± 0.49) | 0.79 (± 0.31) | **0.42 (± 0.24)** |
| flowSOM | 0.04 (± 0.12) | 0.0 (± 0.0) | 0.0 (± 0.0) | 0.13 (± 0.21) | 0.01 (± 0.02) | 0.33 (± 0.4) | 0.17 (± 0.34) | 0.0 (± 0.0) | 1.0 (± 0.0) | **0.19 (± 0.12)** |
| FLOCK | 0.04 (± 0.12) | 0.0 (± 0.0) | 0.0 (± 0.0) | 0.01 (± 0.02) | 0.0 (± 0.0) | 0.31 (± 0.39) | 0.16 (± 0.32) | 0.0 (± 0.0) | 0.77 (± 0.28) | **0.14 (± 0.13)** |
| Rclusterpp | 0.14 (± 0.13) | 0.0 (± 0.0) | 0.0 (± 0.0) | 0.17 (± 0.2) | 0.01 (± 0.01) | 0.35 (± 0.43) | 0.18 (± 0.37) | 0.0 (± 0.01) | 0.95 (± 0.14) | **0.2 (± 0.14)** |
| **Mean: Clustering method** | **0.48 (± 0.16)** | **0.01 (± 0.02)** | **0.17 (± 0.16)** | **0.42 (± 0.23)** | **0.17 (± 0.12)** | **0.49 (± 0.41)** | **0.29 (± 0.37)** | **0.27 (± 0.23)** | **0.93 (± 0.11)** | **0.36 (± 0.2)** |
| **Mean: Classification algorithm** | **1.0 (± 0.0)** | **0.75 (± 0.13)** | **0.98 (± 0.03)** | **1.0 (± 0.0)** | **0.98 (± 0.01)** | **0.85 (± 0.13)** | **0.9 (± 0.12)** | **0.99 (± 0.01)** | **1.0 (± 0.0)** | **0.94 (± 0.05)** |
| **Mean: All** | **0.72 (± 0.08)** | **0.36 (± 0.07)** | **0.55 (± 0.1)** | **0.69 (± 0.12)** | **0.55 (± 0.07)** | **0.66 (± 0.28)** | **0.57 (± 0.25)** | **0.61 (± 0.12)** | **0.96 (± 0.06)** | **0.63 (± 0.13)** |

**Supplementary Fig 1.**
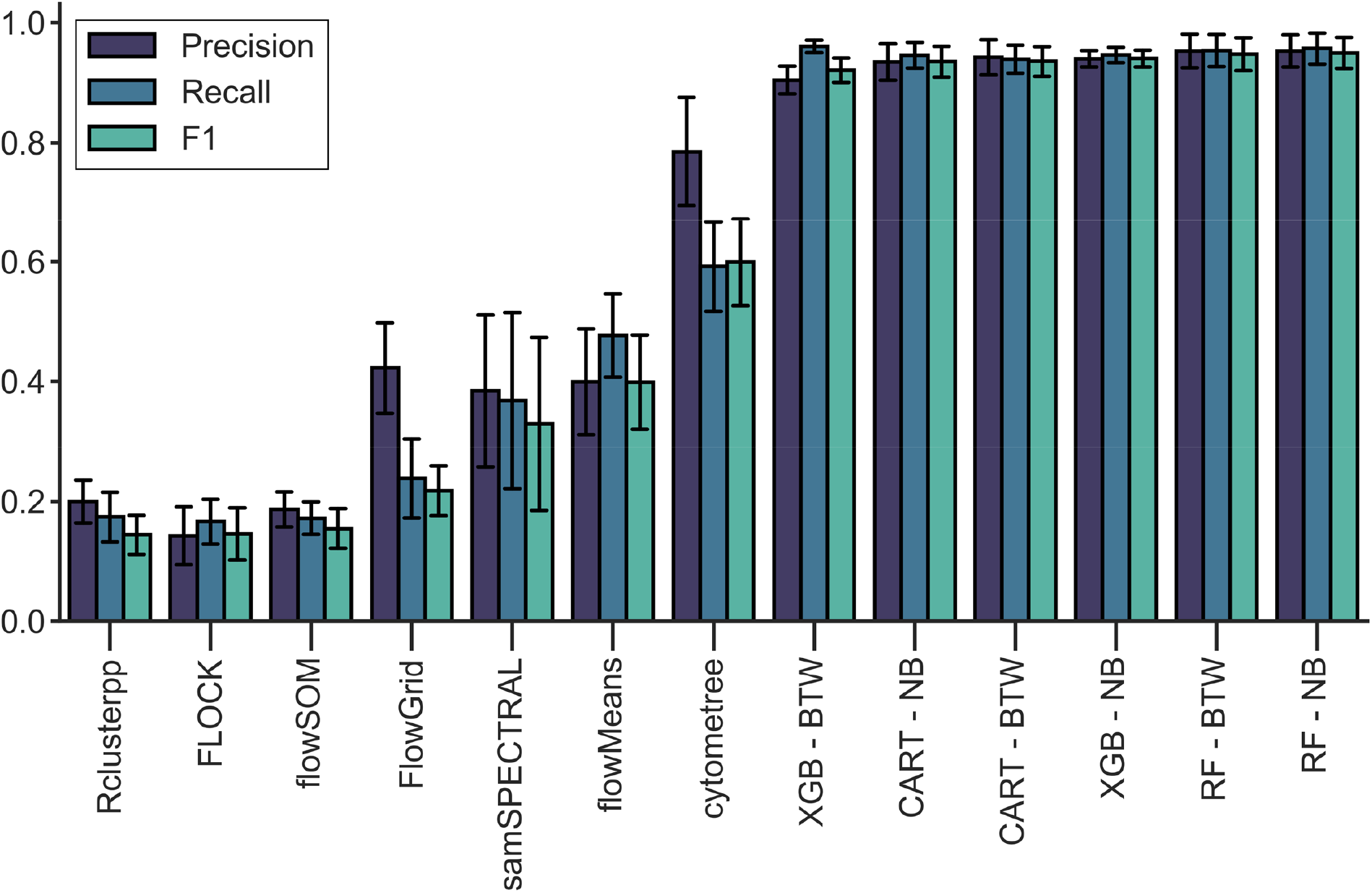
Mean (± SD) precision, recall and F1 for classification algorithms and clustering methods. CART, Random Forest (RF) and XGBoost (XGB) were trained once with models weighting classes inversely proportional to their frequencies within the dataset during model training (Balanced Training Weights = BTW), and once without (No Balancing = NB). Classification algorithms were cross validated over 10 folds with a labelled dataset containing events from 152 samples. Clustering methods were run on 10 randomly chosen samples from the labelled dataset, identified clusters were matched to labelled populations using the Hungarian assignment algorithm.

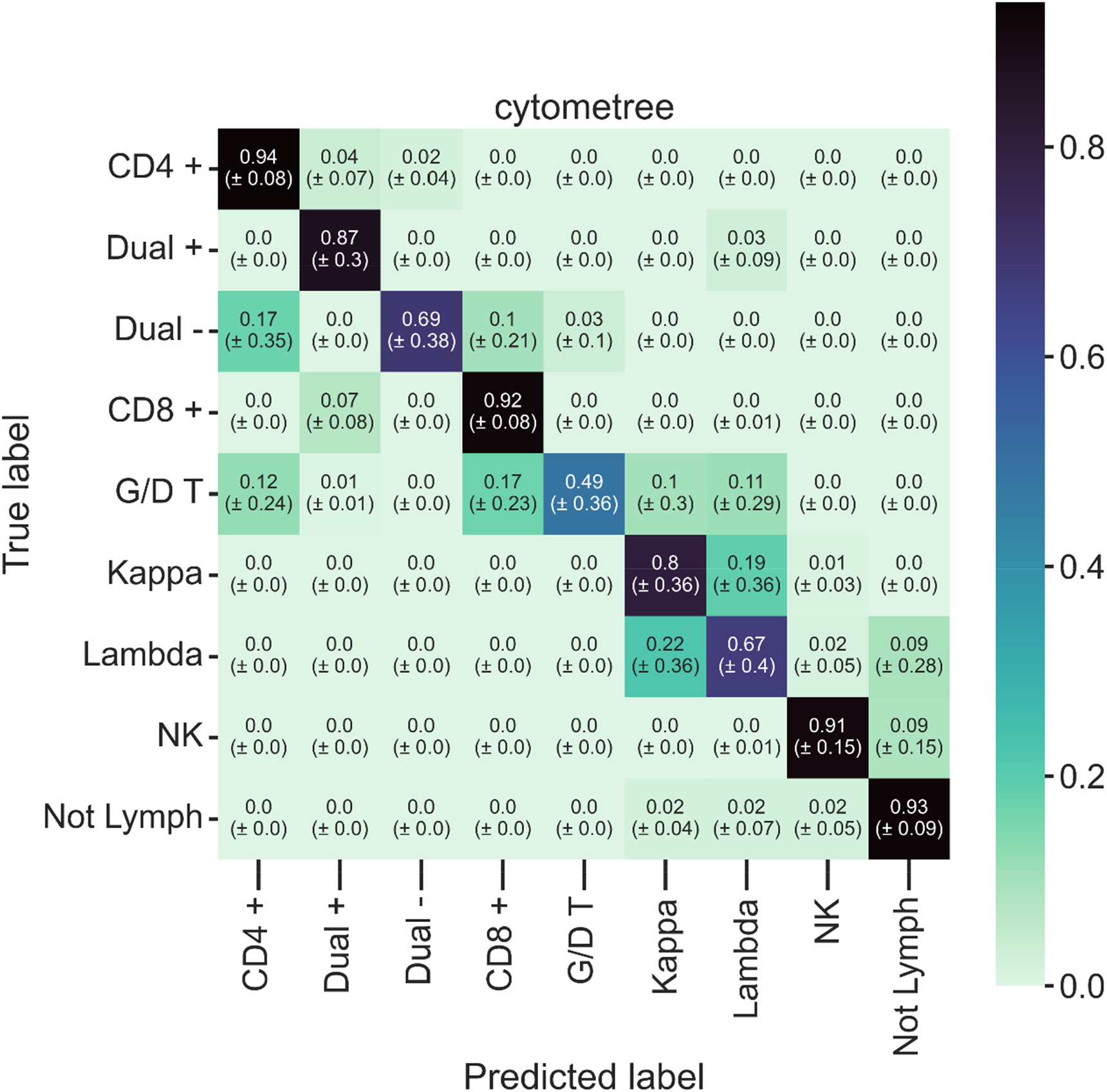

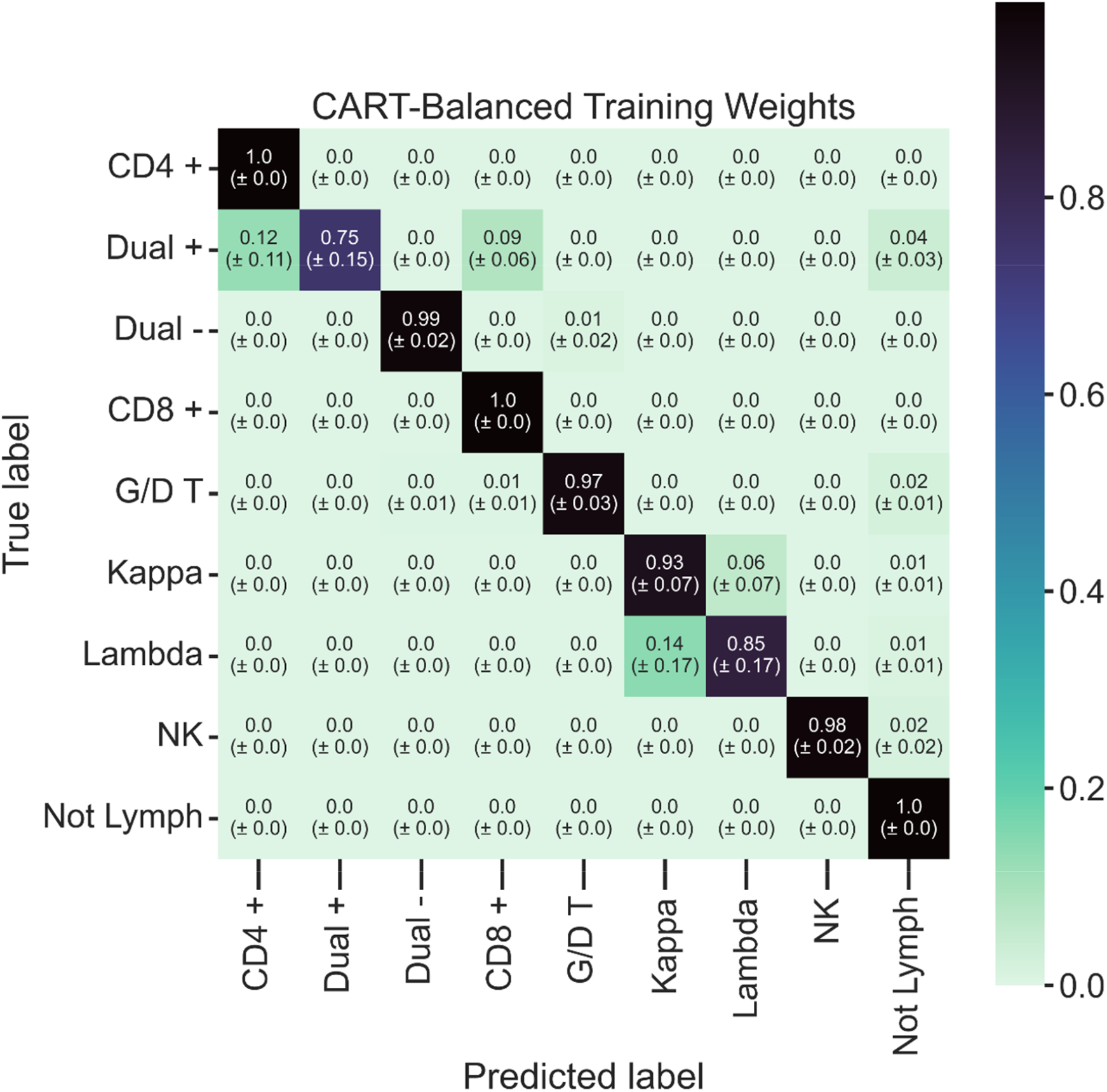

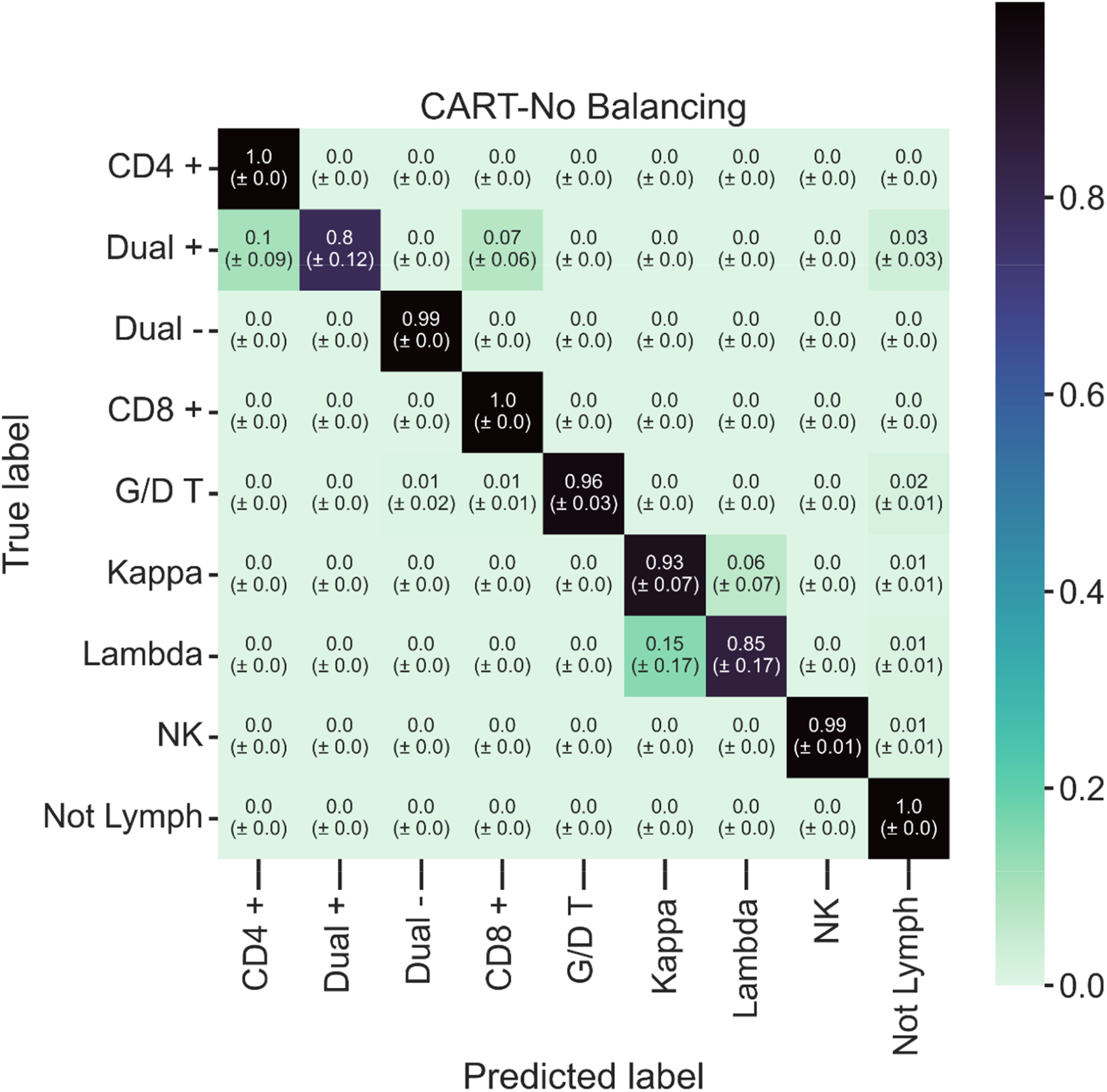

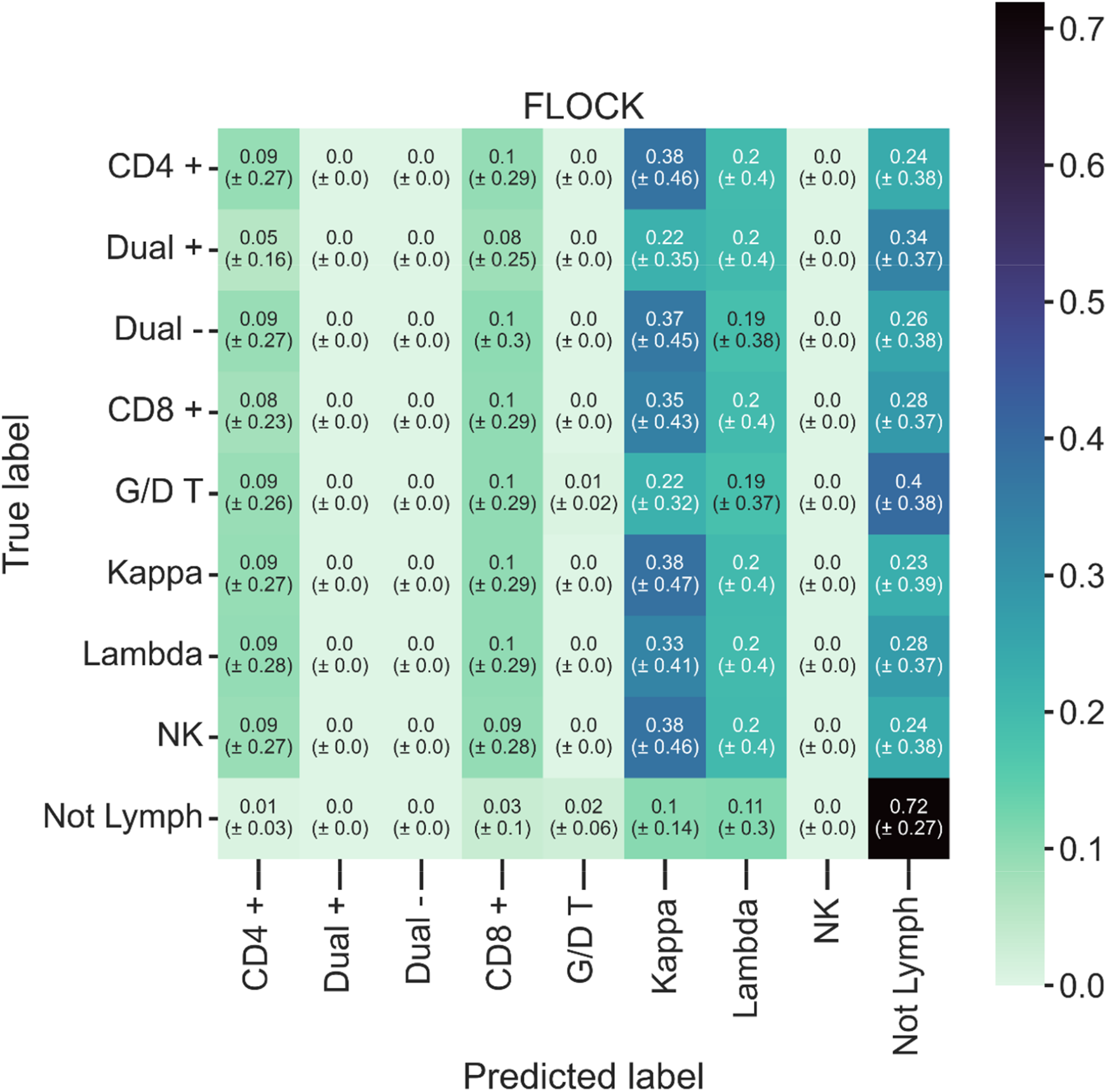

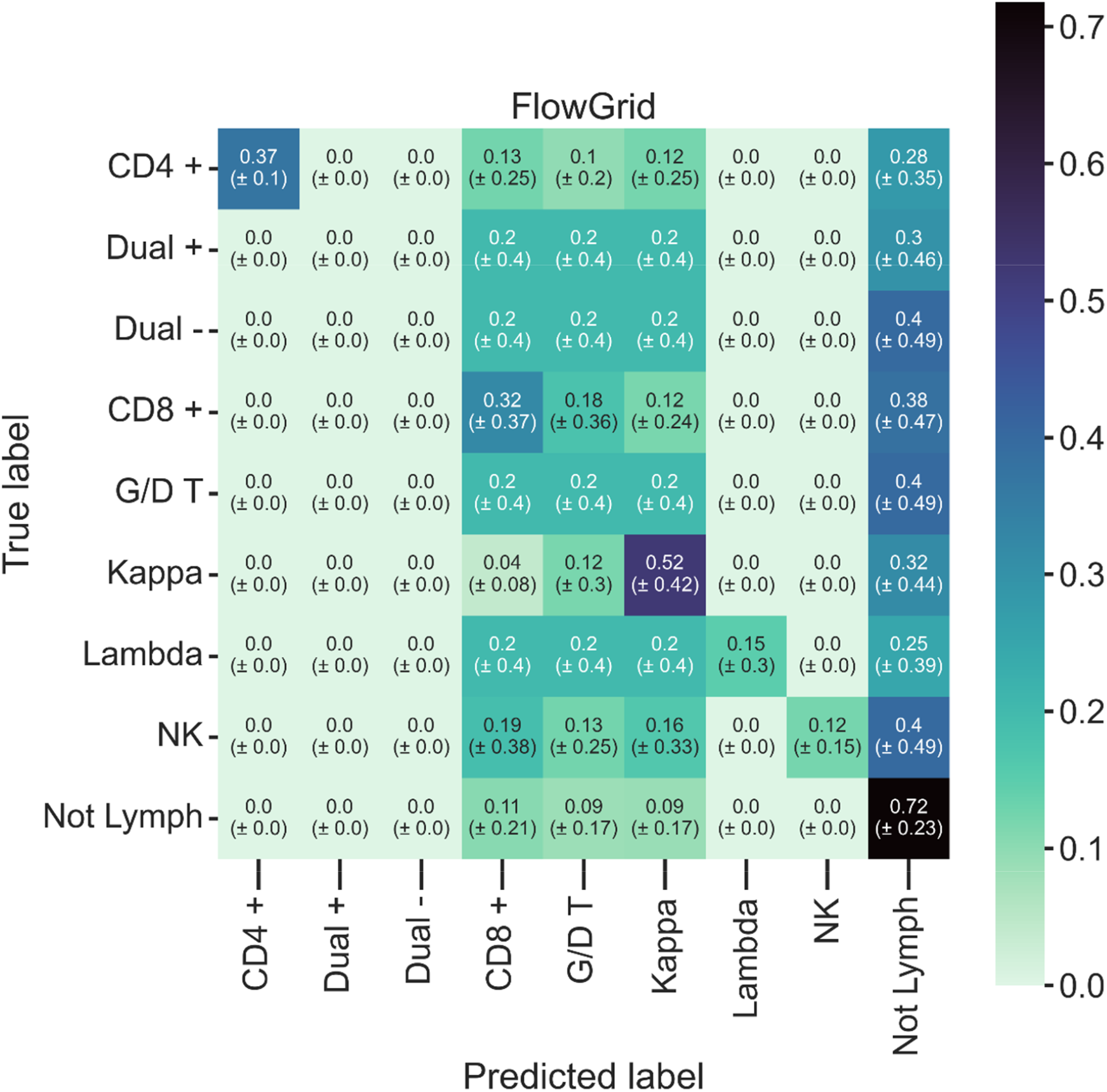

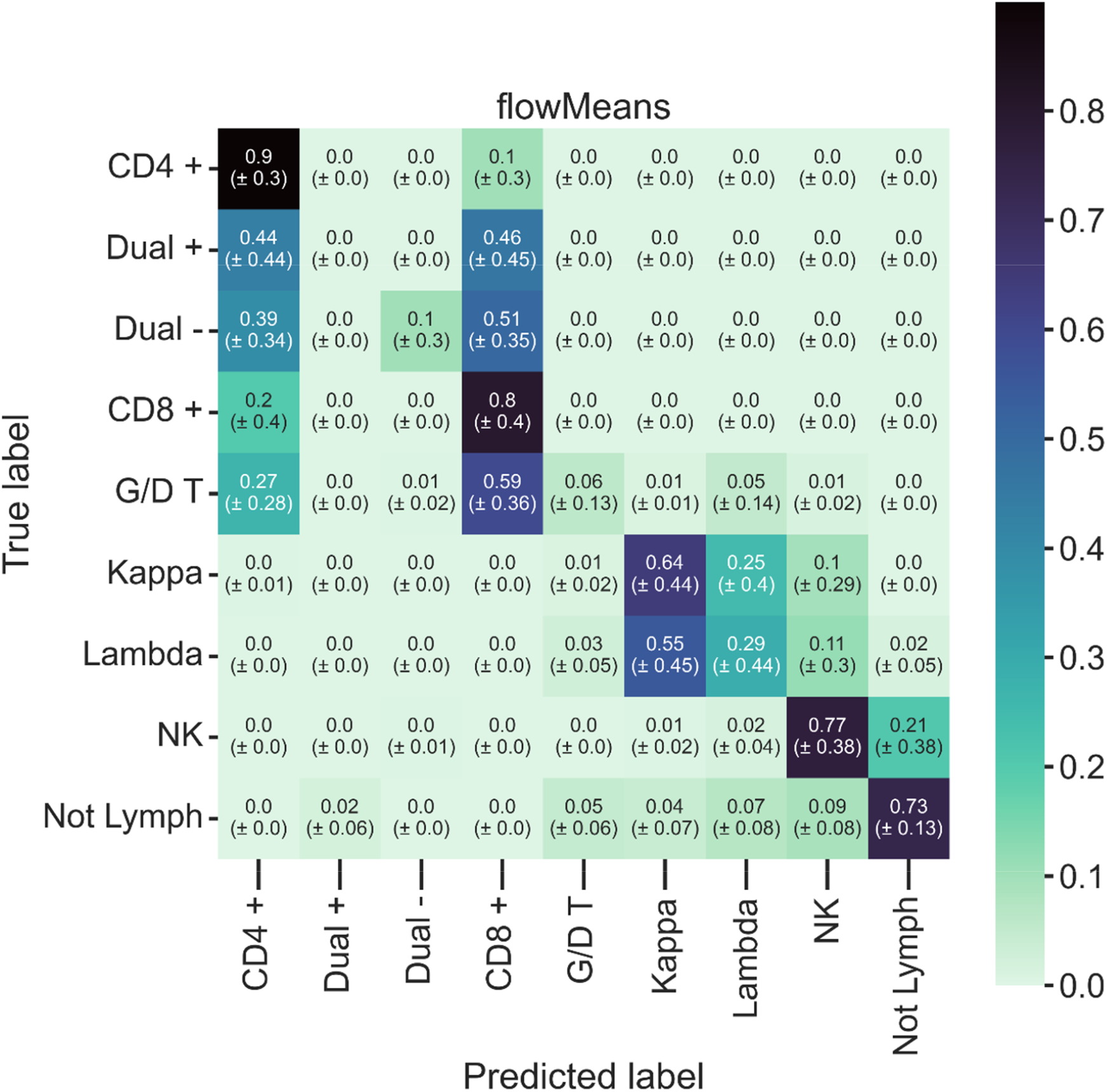

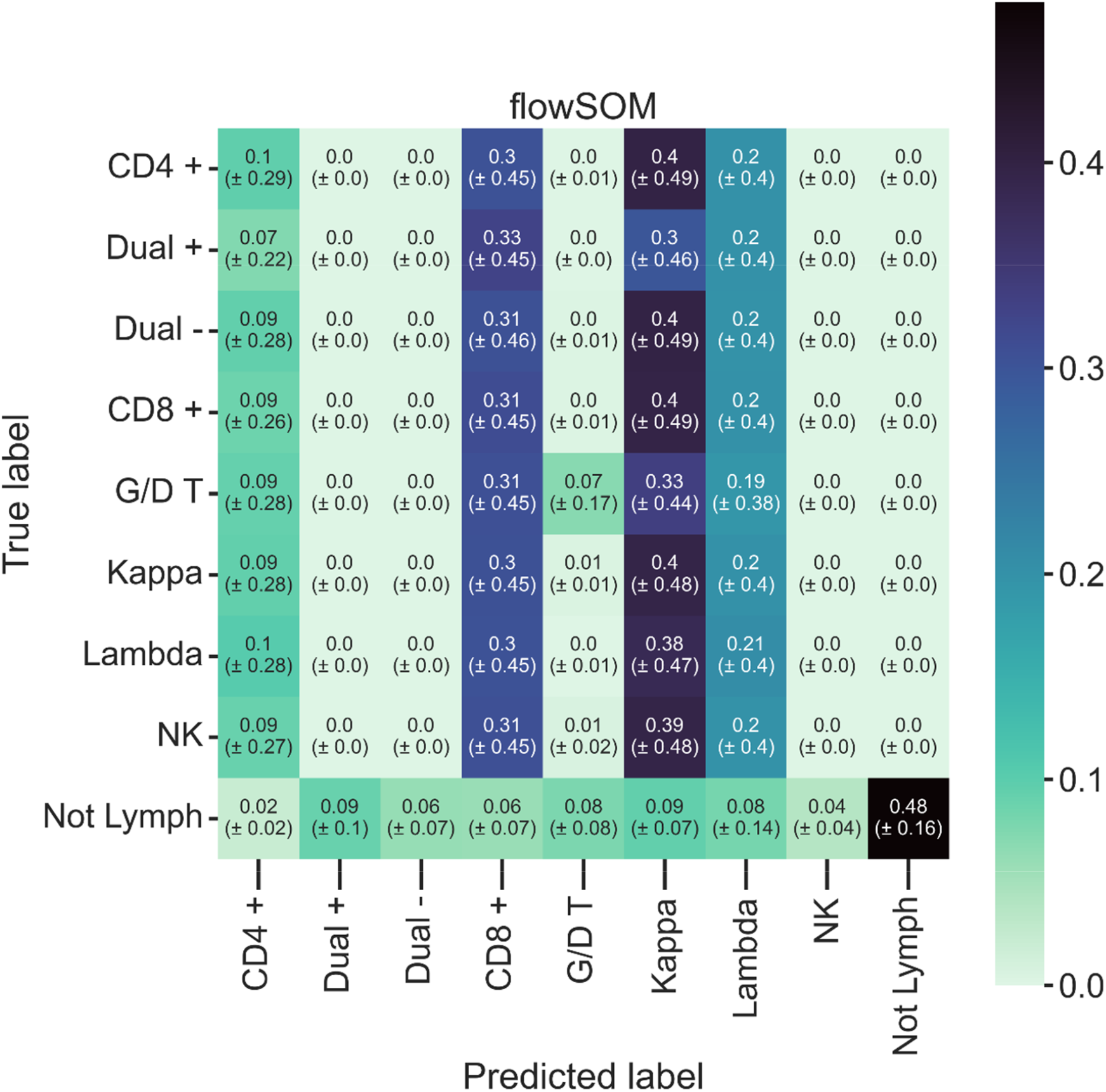

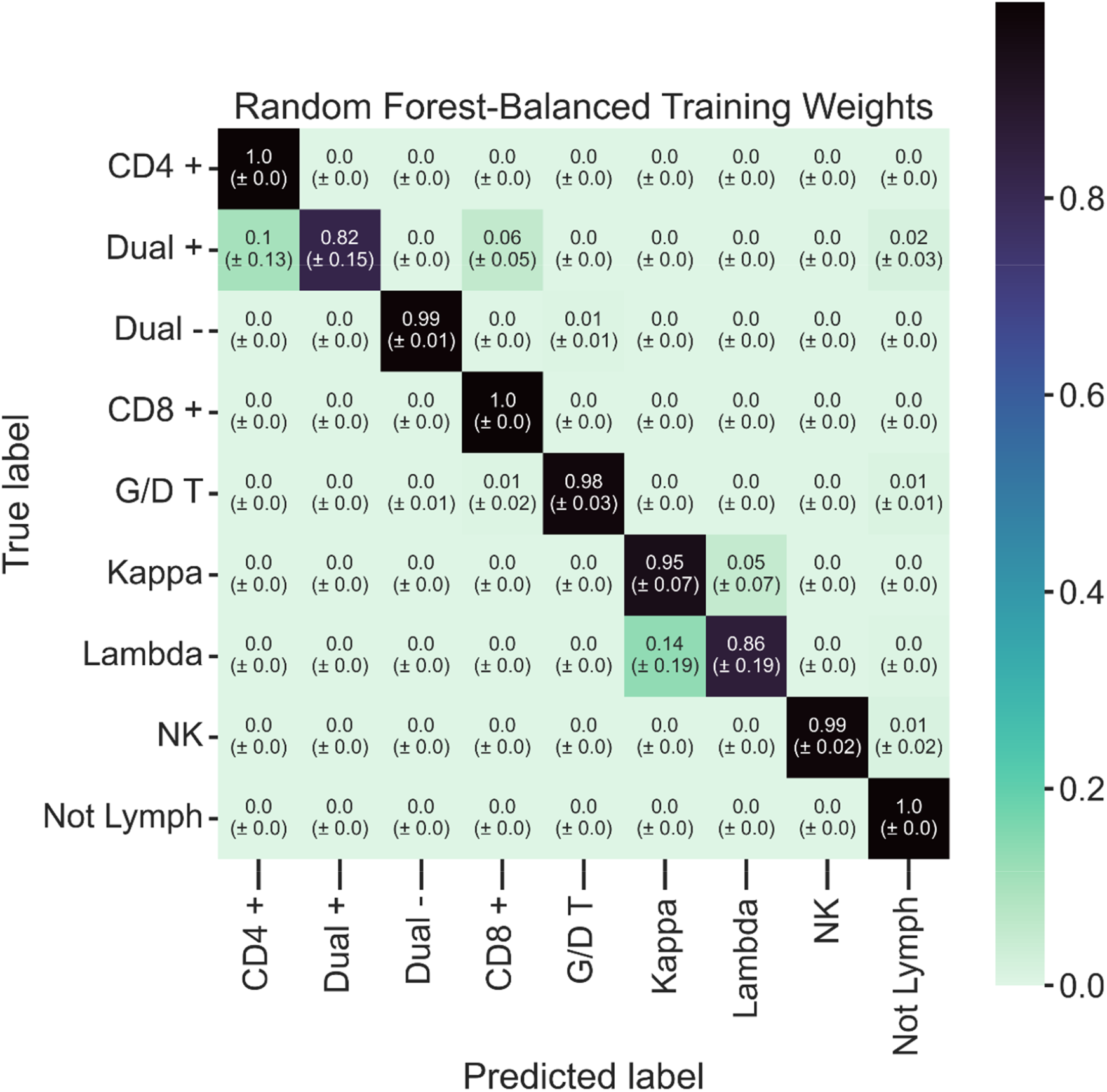

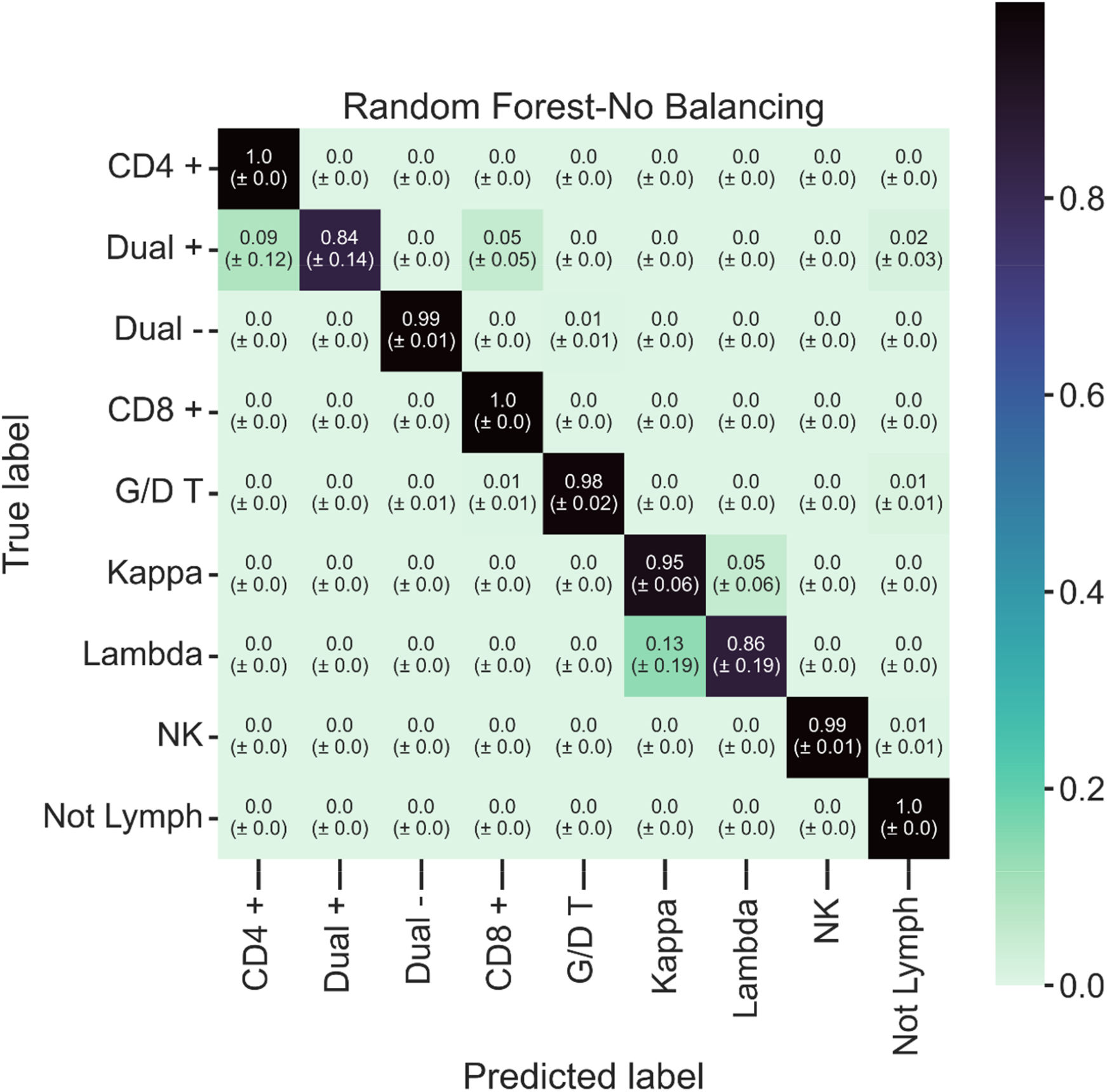

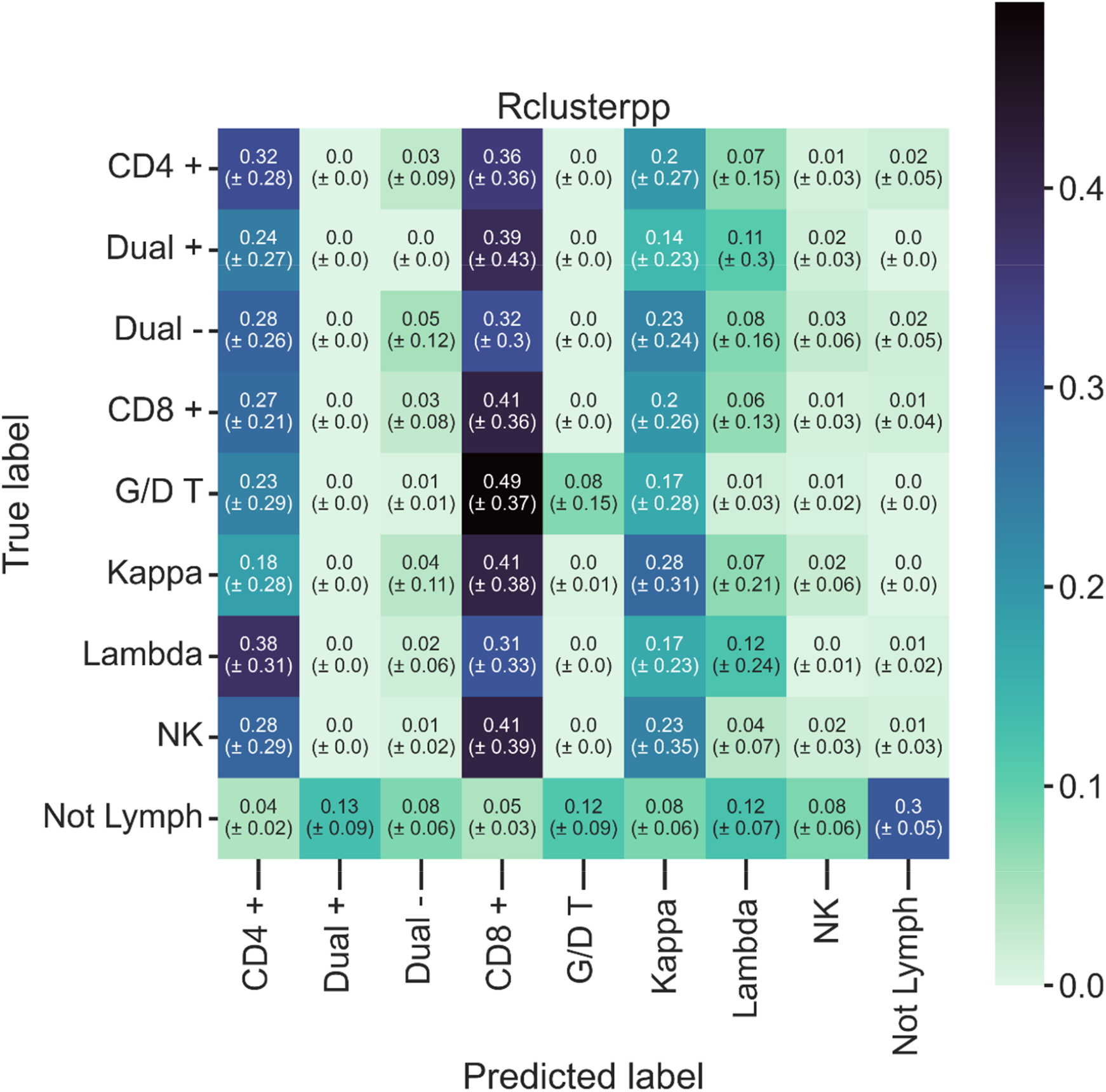

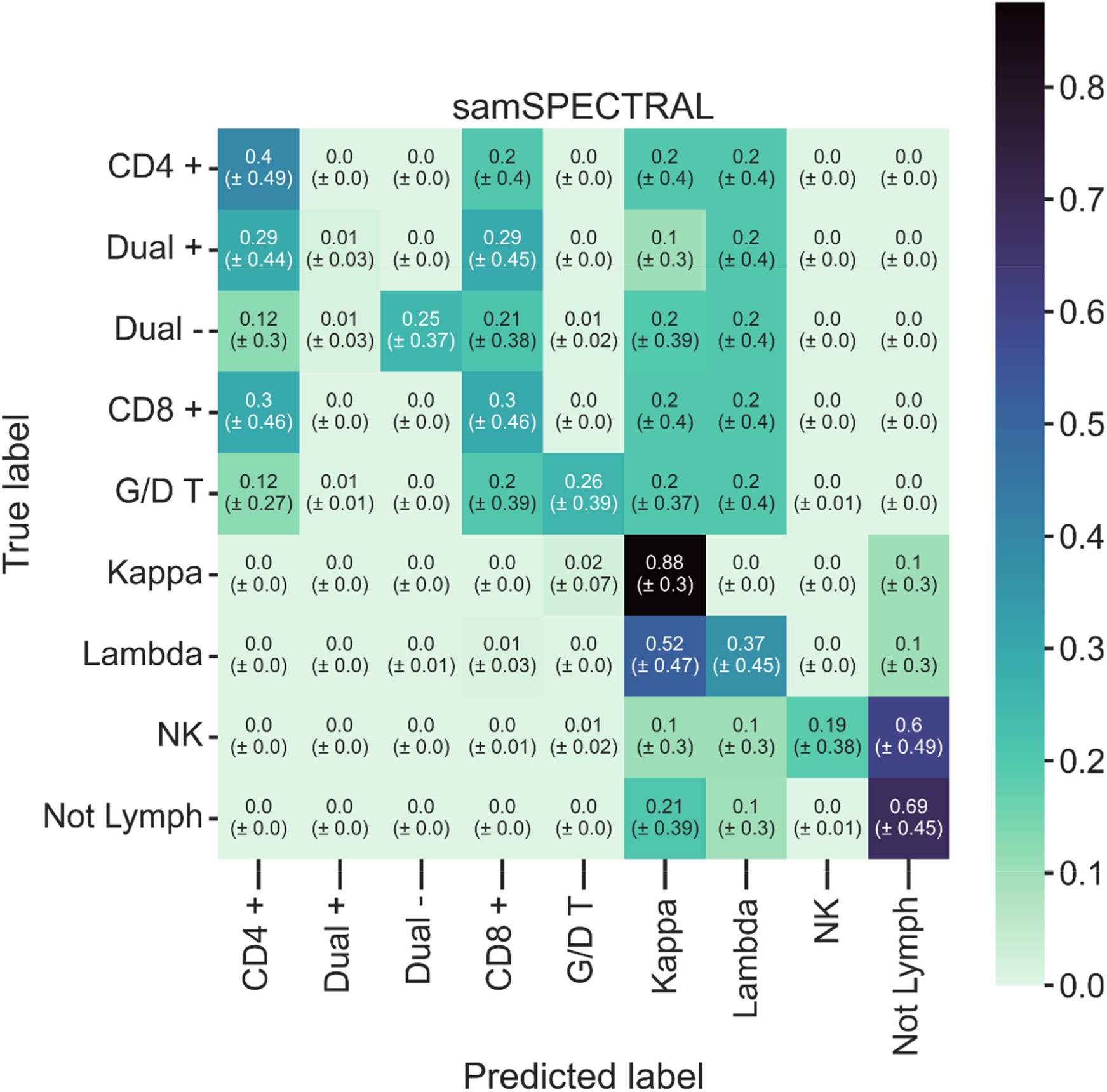

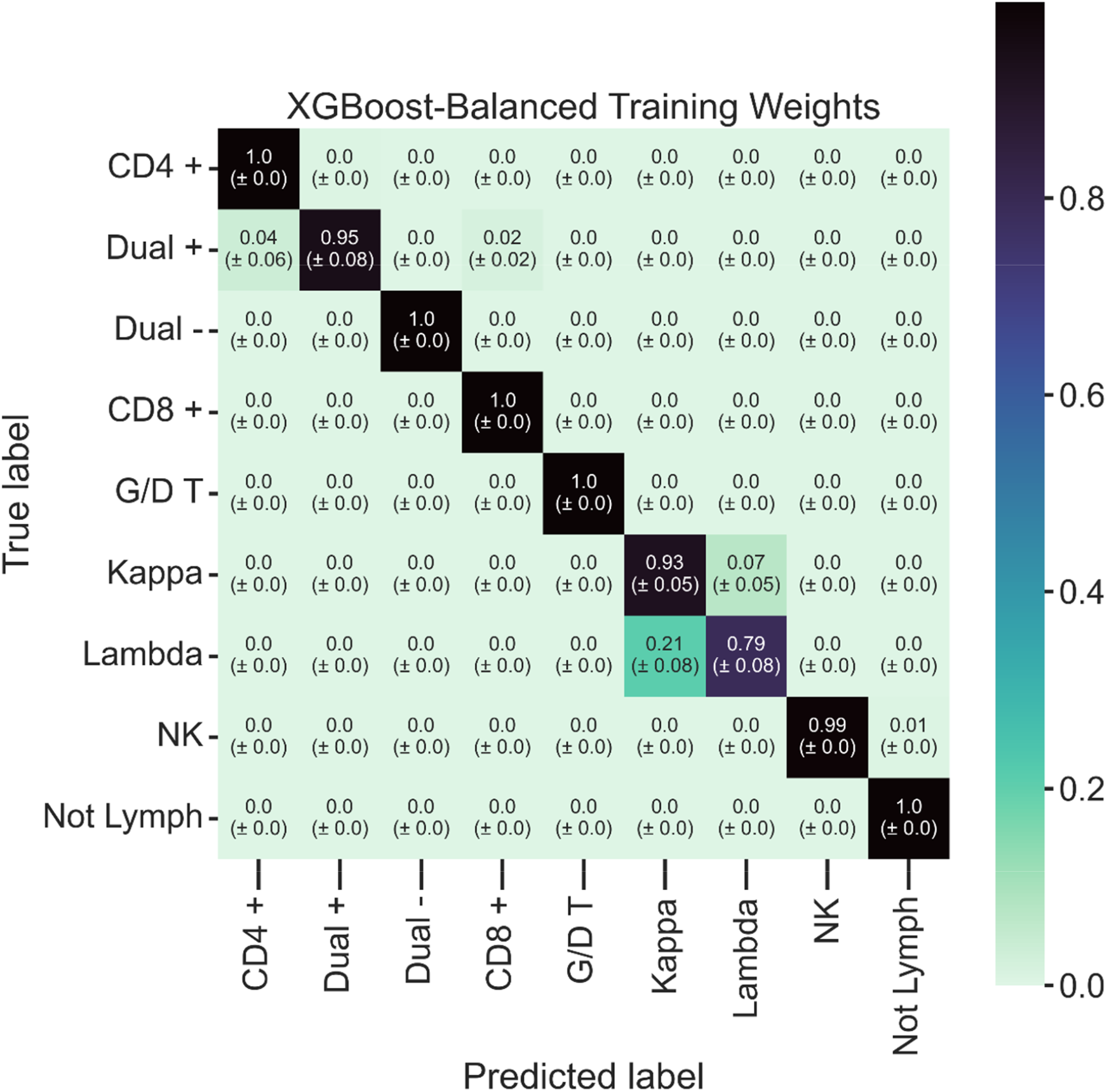

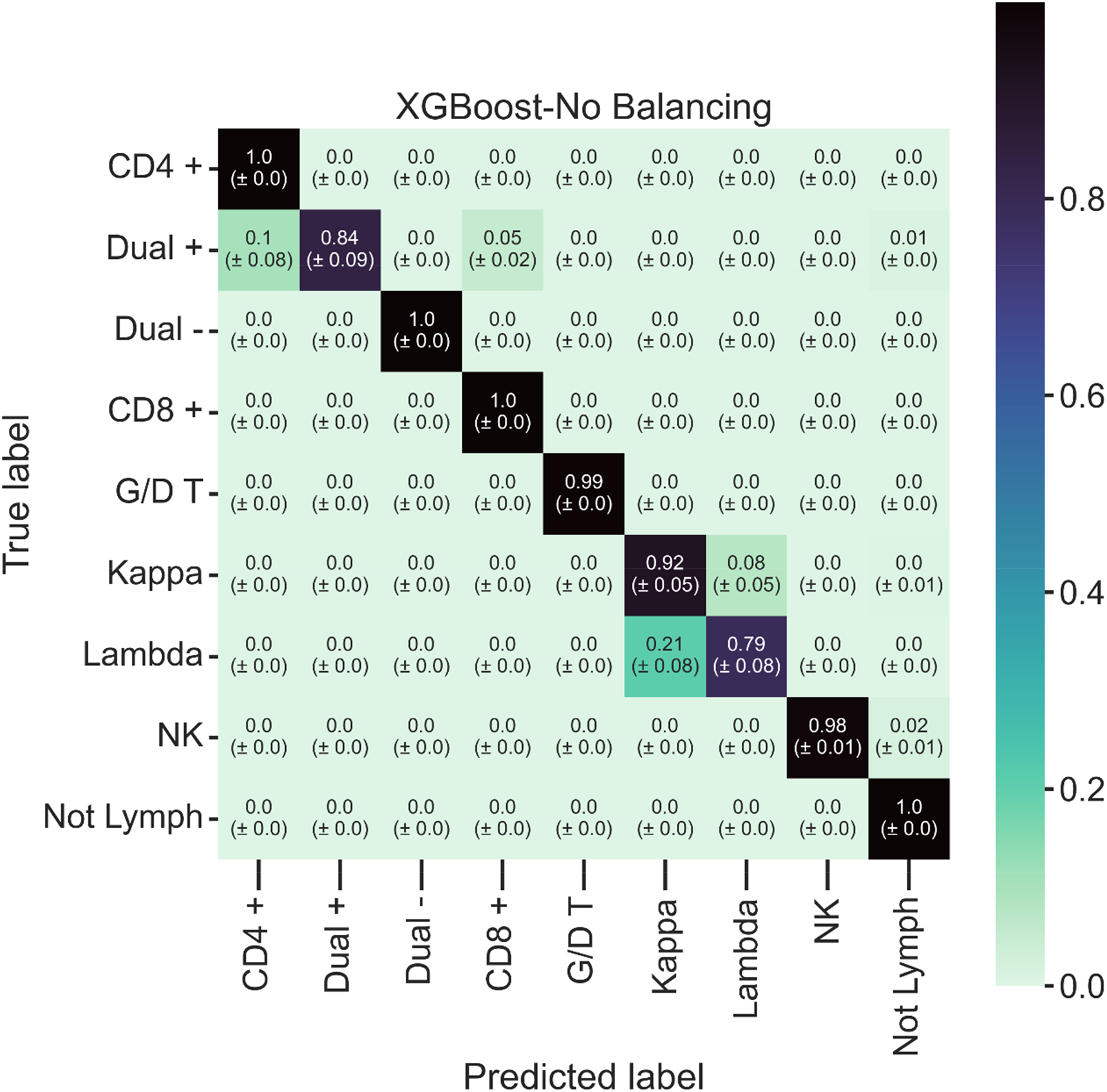

